# Contaminated Sites and Indigenous Peoples in Canada and the United States: A Scoping Review

**DOI:** 10.1101/2022.08.08.22278551

**Authors:** Katherine Chong, Niladri Basu

## Abstract

**Background:** Indigenous communities in Canada and the US are disproportionately exposed to contaminated sites, often arising from industrial and waste disposal activities. For instance, ∼34% of US EPA Superfund sites are of Native American interest, and ∼29% of Canadian federal contaminated sites are on Indigenous reserve land. Contaminated sites pose unique challenges to many Indigenous peoples who consider the land as an integral part of food systems, culture, and the economy. Federal management of contaminated sites is challenged by epistemological differences, regulatory barriers, and minimal scientific research.

**Objectives:** This scoping review aimed to identify and map information on contaminated sites and Indigenous peoples in Canada and the US, namely: 1) the relationship between contaminated sites and Indigenous people, and their land and food systems; 2) strategies, challenges, and successes for contaminated sites assessment and management on Indigenous land; and 3) Indigenous leadership and inclusion in contaminated site assessment and management.

**Methods:** Three streams of data were retrieved from January to March 2022: a systematic literature search (key word groups: Indigenous people and contaminated sites); a grey literature search; and an analysis of federal contaminated site data (Canada’s Federal Contaminated Sites Inventory (FCSI) and US EPA’s Superfund Database).

**Results:** Our search yielded 49 peer-reviewed articles, 20 pieces of grey literature, and 8114 federal site records (1236 Superfund, 6878 FCSI), evidencing the contamination of the lands of 815 distinct Indigenous tribes and nations and the presence of 440 different contaminants or contaminant groups. Minimal information is available on the potential health and ecological effects, assessment and management of risks, and collaboration on contaminated site processes relative to the number of sites on or adjacent to Indigenous lands.

**Discussion:** By integrating three diverse data streams we discovered a multi-disciplinary yet disparate body of information. The results point to a need to prioritize holism, efficiency, and Indigenous leadership in contaminated site assessment, management, and research. This should include a focus on community-specific approaches to site assessment and management; a re-conceptualization of risks related to sites that privileges Indigenous epistemologies; greater collaboration between networks such as the scientific community, Indigenous communities, and federal governments; and a re-evaluation of current management frameworks with Indigenous leadership at the forefront.

## 1. INTRODUCTION

It is well known that pollution has profound effects on human and environmental health (Landrigan et al., 2018). In Canada and the United States (US), contaminated sites that arise from commercial, industrial, or waste disposal activities are a major source of pollution, with contaminant concentrations that can often pose a human or environmental health hazard (Contaminated Sites Management Working Group, 2000; EPA, 2021c). Federal Contaminated Sites (also termed “Superfund” or “Cleanups” in the US) are those which federal governments are responsible for. These sites disproportionately affect Indigenous People. For example, in Canada south of the 60^th^ parallel, there are an estimated 4486 Federal contaminated sites on Indigenous reserve land, making up over 20% of the total sites, although reserves make up only 0.5% of the total land mass (Government of Canada, 2021; OECD, 2020). Similarly in the US, ∼34% of Superfund sites are categorized under ‘Native American Interest’ by the Environmental Protection Agency (EPA), while Native American people make up only 2.9% of the total population (EPA, 2021a; Jones, 2021).

Many Indigenous peoples in Canada and the US have traditional ties to the land, including as an integral part of food systems, language, culture, community, and spirituality. The health of Indigenous communities cannot be understood independent of the health of the environment and thus contaminated sites present unique challenges to Indigenous People (Hoover, 2013). One of the most notable challenges is the impact of contaminants on Indigenous food systems, including agricultural and subsistence activities such as hunting, gathering, farming, and gardening (Fernández-Llamazares et al., 2020). As an example, the First Nations Food and Nutrition Study in Canada recently sampled traditional foods from 92 communities, measuring 2061 samples and finding many of these to have elevated concentrations of mercury, lead, cadmium, and arsenic (Chan, Singh, et al., 2021). Exposure to such contaminants is associated with organ damage, neurodegenerative diseases, cancer, and reproductive and developmental disorders (Briffa et al., 2020; Engwa et al., 2019). Furthermore, contamination of traditional foods has contributed to a ‘nutrition transition’ for many Indigenous communities, characterized by the westernization of diet and lifestyle and a reliance on nutrient-poor market foods. This transition is fueling a high prevalence of chronic diseases including obesity and related cardiometabolic disorders (Chan, Fediuk, et al., 2021; Damman et al., 2008) Overall, exposure to contaminants results in both direct and indirect health effects for Indigenous peoples, contributing to a loss of food sovereignty, which is defined as the right to healthy, culturally appropriate, and self-determined food and agricultural systems, and is a key determinant of overall health (Coté, 2016; Fernández-Llamazares et al., 2020).

Unique solutions are required to address the challenges that contaminated sites pose to Indigenous communities, however environmental research and management processes often use western-institutionalized risk assessment tools and frameworks which are generic and poorly apply to Indigenous people (Arsenault et al., 2019; J. Sandlos & A. Keeling, 2016; Wang et al., 2020). For example, the application of a generic risk assessment model for mercury exposure to studies involving Indigenous populations did not yield results that were representative of the study population (Canuel et al., 2006). This issue is further complicated by the lack of toxicological data for the hundreds of chemicals found at contaminated sites (EPA, 2021d; Government of Canada, 2021; Wang et al., 2020). Regulatory barriers further challenge contaminated sites management. The variety of US governmental programs, for example, that categorize and address contaminated sites has resulted in a fragmented tracking system and a poor understanding of the scope of the problem (EPA, 2021c). In Canada, Indigenous reserve land falls under federal jurisdiction, while most environmental management is provincially mandated (Eckert et al., 2020). The Indian Act constitutes federal legislation on reserve management, but fails to mention environmental protection, and Indigenous legal systems are not consistently recognized under the Canadian constitution on the same level as federal and provincial legislation (Eckert et al., 2020; Gunn K, 2021). Taken together, there are scientific gaps, regulatory barriers, and a lack of meaningful inclusion of Indigenous communities, which prevent improvements to the management of federal contaminated sites.

Contaminated sites threaten land-based food systems that are essential to many Indigenous communities’ culture, spirituality, and overall health. Scientific, regulatory and epistemological barriers facing federal contaminated site management adds complexity beyond previous studies of pollution and Indigenous people in general (Fernández-Llamazares et al., 2020). However, to our knowledge, the topic of federal contaminated sites and Indigenous people has yet to be reviewed, preventing the advancement of solutions to these barriers. As such, the objective was this paper was to present the state of knowledge on 1) the relationship between contaminated sites and Indigenous people, and their land and food systems; 2) strategies, challenges, and successes for contaminated sites management on Indigenous land; and 3) Indigenous leadership and inclusion in contaminated sites management activities. In doing so, this scoping review maps the available information and identifies evidence gaps and priority actions pertaining to contaminated sites and Indigenous peoples in Canada and the US (Munn et al., 2018).

## 2. METHODS

### 2.1 Positionality Statement

This review was conducted by non-Indigenous researchers at McGill University in Montreal, Canada. The authors are non-Indigenous persons of color (a doctoral student and Professor). While every attempt was made to minimize biases, we acknowledge that this positionality and its historical context places limitations on the present work. The motivation for the review is a community-based project lead by the Kanien’kehà:ka (Mohawk) Community of Kanesatake in Quebec, Canada, that addresses a local contaminated site. The authors are based in McGill’s Centre for Indigenous Nutrition and Environment (CINE), which affords connections with many Indigenous communities and stakeholders and provides opportunities for the promotion of Indigenous methodologies and decolonizing research practices.

### 2.2 Search Strategy and Study Selection Criteria

The search strategy utilized three data streams, which included a systematic scholarly literature search, a grey literature search, and federal contaminated sites data repositories (Figure 1). First, a systematic search of peer-reviewed literature was conducted to identify the state of knowledge pertaining to the study objectives. Second, a search for grey literature, such as governmental reports, theses, news articles, and case studies was used to support the findings. Finally, publicly available federal contaminated site data was downloaded from the US and Canadian federal government websites. The results were compiled and scrutinized to map the existing evidence and to develop an understanding of priority scientific and environmental management gaps in a multidisciplinary context. The PRISMA (Transparent Reporting of Systematic Reviews and Meta-Analyses) checklist was utilized to ensure that methodology reporting was transparent (Tricco et al., 2018).

**Figure 1:**
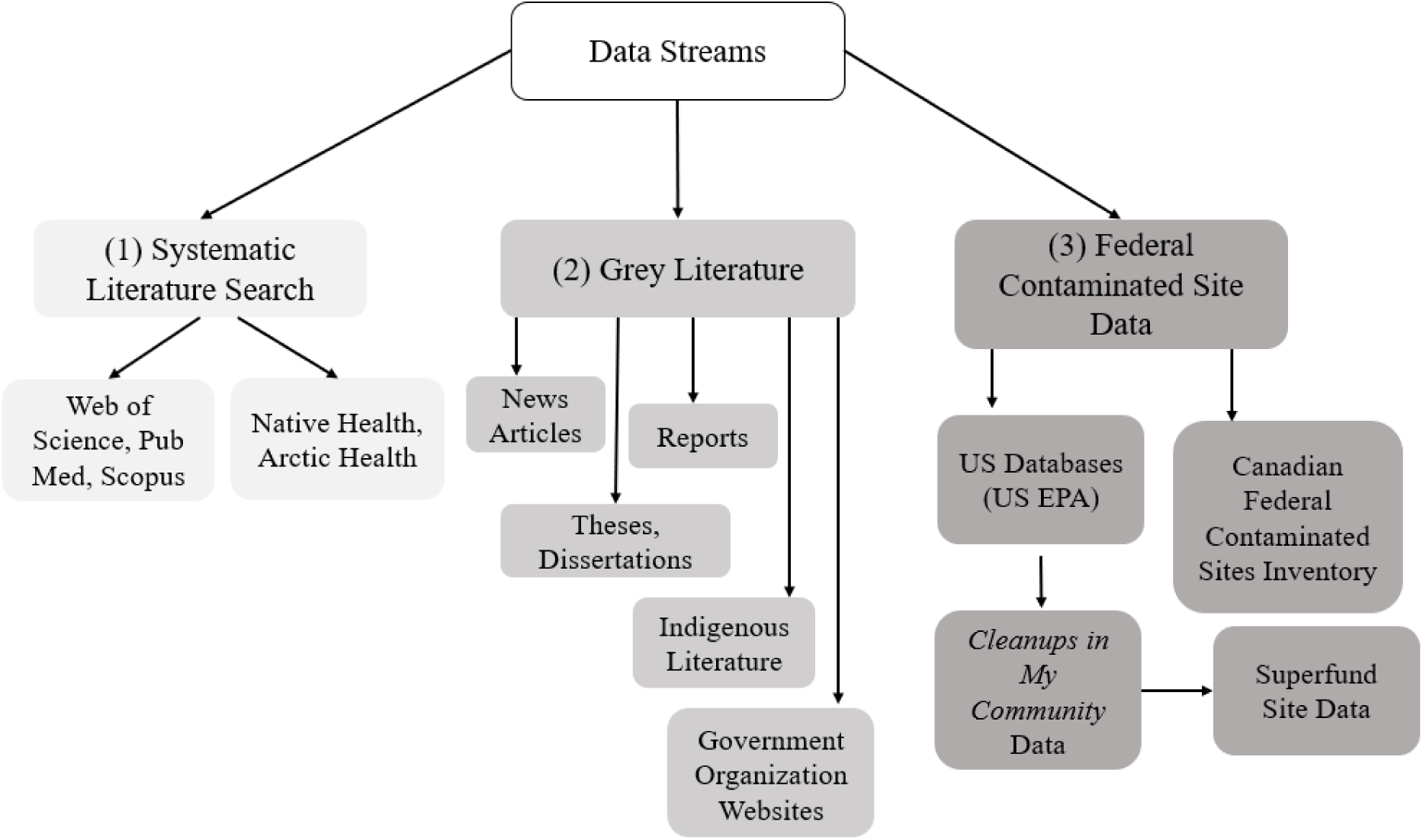
Overview of the three data streams used in this scoping review

#### 2.2.1 Systematic Literature Search

A systematic search of databases (Scopus, Web of Science, and Pubmed) was used to identify relevant articles. The final search was run on January 10, 2022. Boolean operators were used to search each database as follows: (“First Nations” OR “Indigenous people” OR “Native American” OR “Aboriginal” OR “Inuit” OR “Metis” OR “American Indian” OR “Canadian Indian” OR “Traditional food” OR “Indigenous agriculture” OR “Country food*” OR “Tribal land” OR “reservation” OR “reserve land” OR “Indian country”) AND (“contaminated site” OR “superfund” OR “contaminated land” OR “brownfield” OR “abandoned mine”). The Scopus search was limited with the operator TI-ABS-KEY due to initial results from a full-text search that lacked specificity.

Overall, inclusion criteria were broad, to capture the diversity and scope of the information available on the topic. Papers were included that pertained to federal contaminated site(s) in Canada or the US, and to Indigenous peoples, communities, and environments exposed to the site. Both qualitative and quantitative studies were included. Both primary studies (i.e., involving the collection of primary data and direct measurement of an outcome of interest) and secondary studies (i.e., review papers or discussion papers) were included. Literature included was limited to Canada and the US and published in English. No date limit was placed on the studies. The review was limited to First Nations, Inuit, and Metis peoples in Canada, and all Native American peoples in the US, including those residing in non-contiguous states. Notably, many mainland Indigenous people such as the Haudenosaunee, Cree, and Anishinaabe live in both the US and Canada, due to the cutting of common cultural territory by the international border (*Native Land Interactive Map*, 2021). Furthermore, some Indigenous communities live on both sides of the US-Mexico Border, and these were considered if the literature was relevant to the US context. The review used the definition of ‘contaminated site’ as described by the US EPA and Environment and Climate Change Canada (ECCC) (See Supplemental Materials, Glossary of Key Terms) (EPA, 2021c; Government of Canada, 2021). The review excluded literature on pollution and Indigenous people that was not specific to contaminated sites. The search and screening process, involving the reviewing of titles and abstracts, resulted in a total of 49 articles for analysis (Figure 2; Excel Table S1).

**Figure 2:**
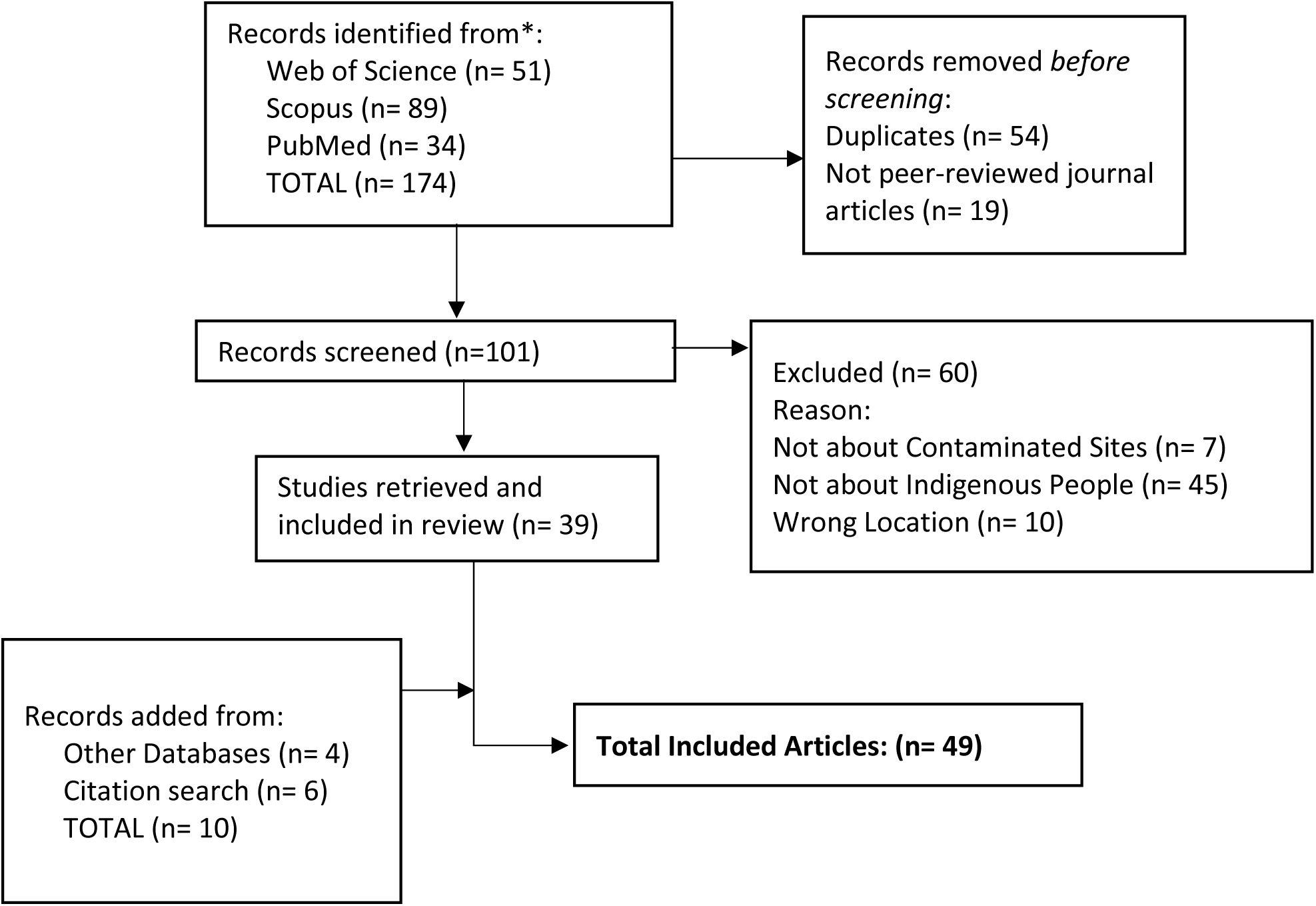
Adapted PRISMA Chart for Scoping Review for first data stream (systematic literature search) (Tricco et al., 2018)

#### 2.2.2 Grey Literature Search

Due to the minimal number of publications in academic journals related to contaminated sites and Indigenous People in Canada and the US, and to capture a broader range of perspectives on the topic, a grey literature search was also conducted. Grey literature included multiple document types produced by all levels of government, academics, businesses and organizations, that are not controlled by commercial publishing. (Mahood et al., 2014) The search strategy ‘SYMBALS’ (a systematic review methodology blending active learning and snowballing) (van Haastrecht et al., 2021), which uses backward snowballing to allow researchers to complement their set of relevant papers with additional sources such as grey literature. The grey literature search was conducted from January 2022 through to March 2022 inclusive.

Grey literature was yielded iteratively from relevant websites, governmental databases, organizations, and reference lists. Search terms were compiled from the authors’ knowledge on the topic and information from previous searches. The search terms used fell into two main categories (“Indigenous People” and “Contaminated Sites”) with the complete list available in Supplemental Materials (Table S1). News articles, reports, dissertations and theses were found in ProQuest and included if relevant. Governmental websites, including Health Canada, Indigenous and Northern Affairs Canada, Environment and Climate Change Canada, US EPA and the Tribal Lands Assistance Center were searched. Information on Indigenous peoples and federal contaminated sites is often cited in technical reports and therefore, Google Scholar was searched, which has been cited as an acceptable database for grey literature searches (Haddaway et al., 2015). The search yielded a total of 20 selected grey literature articles (Excel Table S2), which included seven government documents (EPA, 2015; Government of Northwest Territories, 2021; Indigenous and Northern Affairs Canada, 2016; Michelsen, 2010; US Government Accountability Office, 2019; US Government Publishing Office, 2016; Woolford, 2017); four journal articles (non-peer-reviewed) (Castleden et al., 2017; Gallo, 2011; Gover, 2007; Lewis et al., 2015), one opinion article (Nolan, 2009), one thesis (Clark, 2020), three news articles,(Bienkowski, 2012; Hansen, 2018; Indian Country Today, 2013), and four conference materials (Ellison, 2012; Gailus, 2013; Hykin, 2016; Kent, 2016). Although not considered as grey literature in this review, information was also gathered from nine government webpages, to better understand the context of the findings (Table S2).

#### 2.2.3 Federal Data Inventory Search

Government websites (US EPA and Government of Canada) were used to identify available datasets for inclusion. Data on contaminated sites was publicly available in an Excel spreadsheet format for all federal contaminated sites on reserve land in Canada from the Federal Contaminated Sites Inventory (FCSI) (n= 6878) (Government of Canada, 2021) (Excel Table S3), which was downloaded on October 14, 2021. A search for US EPA Superfund sites categorized under “Native American Interest” (which includes sites that affect Native American peoples but are not on Tribal lands) from the Superfund Site Information database was run on October 4, 2021, and re-run on February 16, 2022 to include both the Superfund site data (n= 1236) (Excel Table S4) and an inventory of data on contaminants at Superfund sites (Excel Table S5) (EPA, 2021d). The searches of the Canadian and US databases resulted in a total of 8114 records of contaminated site data for analysis. Canadian and US data was not consistent, however some variables were available in both datasets, which included the site location and Indigenous community affected, site status (i.e., whether the site was currently active), risk classification and prioritization ranking, type of contaminant, and type of contaminant media. Canadian FCSI data also included information on the management strategy and plan for the site, the estimated size of the area contaminated, the number of people living in proximity to the site, and expenditure estimates related to monitoring, remediation, assessment, and maintenance. Superfund data included indications of whether the contaminants were of human or ecological concern, the site type (i.e., source of contamination), and exposure control measures.

### 2.3 Data Extraction and Analysis

Included peer-reviewed literature were compiled into a data chart following a model outlined by the Joanna Briggs Institute Methodology for Scoping Reviews (Peters et al., 2015) (Excel Table S1). Per this guideline, the following data were extracted and charted: author(s), year of publication, country of origin, study purpose, location, study population (i.e., sample size), methodology, and key findings related to the scoping review questions. Additionally, data extracted that are specific to this study included: contaminated site name, contaminant name or contaminant class, contaminant media sampled, type (source) of contaminated site, Indigenous community of focus and/or territory of focus, the presence of a collaboration between Indigenous and non-indigenous researchers and/or Indigenous authorship, whether the article was a primary or secondary data source, whether the article collected qualitative or quantitative data, and the discipline in which the article was published. For quantitative data, descriptive statistics were used to map out and better understand the scope of the data. Qualitative data were iteratively analyzed to develop themes presented in the results section.

Data from both retrieved articles and federal databases were analyzed using Microsoft Excel Software (Microsoft 365 MSO (Version 2205)), and basic descriptive statistical analyses were run on these datasets. For example, data such as contaminants of focus, risk assessment information, and site locations were extracted and compared across data streams. Data was analyzed in answer to the three study objectives, with additional questions being developed iteratively as the data was explored. A narrative synthesis of qualitative data was integrated into this analysis. Complete datasets can be found in supplementary materials, which includes data extraction charts for peer-reviewed literature (Excel Table S1) and grey literature (Excel Table S2); and raw data retrieved from Canadian FCSI (Excel Table S3) and US EPA Superfund databases (Excel Tables S4 and S5).

## 3. RESULTS

### 3.1 Overview of results

Information available on contaminated sites and Indigenous peoples in Canada and the USA included 49 peer-reviewed journal articles, 20 pieces of grey literature, and 8114 contaminated site records from federal databases (Table 1). These sources provided data on the potential or actual contamination of the lands of 815 distinct Indigenous tribes and nations and indicated the presence of 440 different chemicals or chemical groups found at 4976 distinct contaminated sites. Peer-reviewed literature available on the topic has been published between 1996-2021. Of the 49 peer-reviewed articles, 35 were primary data sources (i.e., original data collection), while 14 were secondary, including reviews, historical reviews, and discussion papers. Of the articles collecting primary data, 26 collected quantitative data, three collected both qualitative and quantitative data, and six collected qualitative data only. Throughout these sources, contamination of a wide variety of media, from soil, air, water, groundwater, and food to human biomarkers were described.

**Table 1:** Meta-data from three data streams (systematic literature review, grey literature, and federal contaminated site databases)

|  | *Data Peer-reviewed Literature* | *Grey Literature <sup>a</sup>* | *Canadian Federal Contaminated Sites Inventory (FCSI)* | *US EPA Superfund Dataset* | *TOTAL* |
| --- | --- | --- | --- | --- | --- |
| *Total # of records* | 49 | 20 | 6878 | 1236 | 8180 |
| *# of active sites <sup>b</sup>* | n/a | n/a | 2428 | 463 | 2891 |
| *# of archived sites <sup>c</sup>* | n/a | n/a | 2843 | 773 | 3616 |
| *# of suspected sites <sup>d</sup>* | n/a | n/a | 1607 | n/a | 1607 |
| *Indigenous communities identified <sup>e</sup>* | 41<br>(8 Canada, 30 US, 3 US and Canada) | 11 | 576 | 239 | 815 <sup>f</sup> |
| *# of contaminants identified* | 45 | 14 | 12 | 440 | 440 <sup>f</sup> |

<sup>a</sup> Grey literature was collected iteratively using SYMBALs methodology as described above (van Haastrecht et al., 2021), and therefore is not representative of the entire scope of grey literature available on the topic
<sup>b</sup> “Active Sites”: have not completed the process for addressing a contaminated site and continue to pose an environmental and/or financial liability to the federal government are classified as active (Environment Canada, 2012)
<sup>c</sup> “Closed” or “Archived” sites are defined by federal governments as having shown acceptable levels of human and ecological risk by meeting national guidelines, remedial objectives have been achieved, and no further action is required (Environment Canada, 2012)
<sup>d</sup> A “suspected site” is one at which further assessment is required to determine if the site is ‘contaminated’ (i.e., remedial action may be required) (Government of Canada, 2021)
<sup>e</sup> The literature review’s definition of Indigenous included First Nations, Inuit, and Metis peoples in Canada, and all Native American peoples in the United States, including those residing in non-contiguous states; Canadian FCSI data was filtered by sites located on reserve land, and inventories the name of the Indigenous community that lives there; Superfund data specifies the Native American community which is affected by the site (with the site classified under “Native American Interest”)
<sup>f</sup> duplicates removed

### 3.2 Indigenous Communities and Geographic Patterns

Peer-reviewed literature described contaminated sites affecting 41 distinct Indigenous Tribes and Nations across Canada and the US (Table 1), with some literature focused on multiple Tribes or Nations. A total of 11 individual communities were the focus of grey articles, with the remaining grey literatures focused on Indigenous Peoples in general as opposed to individual communities. In contrast, more communities were captured under the federal databases as the US EPA reported Superfund sites of interest to 239 Indigenous Tribes (EPA, 2021d), and Canada’s FCSI included sites affecting 576 distinct Indigenous communities on 745 different reserve lands (Excel Tables S3 and S4) (Government of Canada, 2021). Notably, Canadian and US federal contaminated site data varied, in that Canada listed multiple “sites” with unique identifiers in the same geographic region, while Superfund tended to list one larger area with a single identifier.

Of the 49 peer-reviewed journal articles that were included in the review, 29 were based in the US, 13 were based in Canada, and seven were based in both the US and Canada. In grey literature, the US EPA published a report in 2020 estimating that 146 605 Native American people live within one mile of a Superfund site, and 449 849 live within three miles. (EPA, 2020) Canadian FCSI data provided an estimate of populations (which included Indigenous and non-Indigenous people) living near contaminated sites on reserves, totaling 2 096 197 people living within 1 km (Government of Canada, 2021). Canadian FCSI data also reported estimated sizes of the contaminated areas, ranging from 0.0001 to 1,500,000 cubic square meters per site (Government of Canada, 2021). Figure 3 displays contaminated site locations identified from the four data streams.

**Figure 3:**
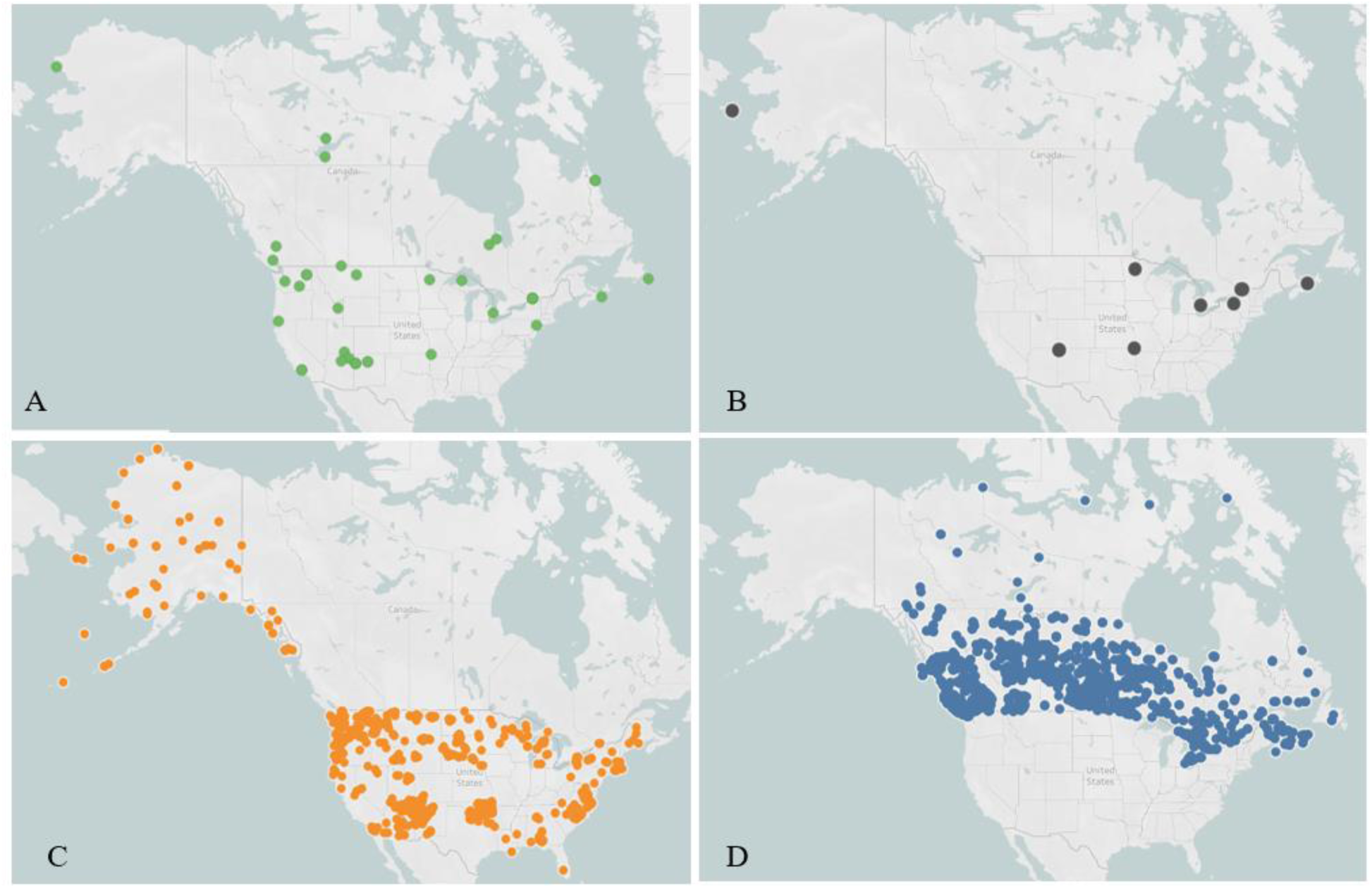
Co-location of Contaminated Sites affecting Indigenous Peoples based on a review of data from A) Site locations identified via the primary literature review; B) Site locations identified in grey literature; C) Site locations identified in US EPA Superfund Database; D) Site locations identified in Canadian Federal Contaminated Site Inventory Data. Each site is indicated with a dot (note: some site locations are the focus of more than one peer-reviewed article). The maps were generated using OpenStreet Maps and Tableau Software. The underlying database is available in the supplementary materials (Excel Tables S1-S4).

### 3.3 Contaminated sites, sources, and media

Of the contaminated sites described in the 49 peer-reviewed articles, 25 were from mining activity, eight were industrial waste sites, six were hazardous waste sites, four were military radar, and one was from hydroelectric activities, with the remainder of unspecified origin. Nine grey articles were focused on specific contaminated sites, which included the mention of seven sites due to mining activities, two from industrial activities, one nuclear waste site, and one from an accidental spill. The US EPA reported on Superfund Site types, including 159 due to manufacturing/processing/maintenance, 84 mining sites, 98 waste management sites, and 39 recycling sites (EPA, 2021d). However, we note that most of the sites (n=248) were classified as having “other” sources (EPA, 2021d). Canadian FCSI Data does not provide information on contaminant source (Government of Canada, 2021).

In the peer-reviewed literature, a total of 45 unique contaminants or contaminant groups were identified or discussed (Table 1). While most grey literature was focused on contaminated sites in general without referencing specific chemicals, seven articles referenced (but did not measure) specific contaminants of concern, which included lead, arsenic, PCBs, benzene, cadmium, formaldehyde, uranium, zinc, pentachlorophenol, dioxin, and creosote. Superfund site data reported the measurement of 440 different contaminants or contaminant groups found at 296 contaminated sites, with lead, arsenic, and cadmium as the most common ones listed. Of these contaminants listed, 12% were flagged by US EPA as a potential ecological risk.(EPA, 2021d) Canadian FCSI Data reported contaminant classes as opposed to single contaminants, for a total of 12 different contaminant classes found at 4680 contaminated sites (Table 1). The most commonly reported classes were petroleum hydrocarbons (PHCs), polycyclic aromatic hydrocarbons (PAHs), and metals, metalloids, and organometallics.(Government of Canada, 2021) In Canadian FCSI Data, there were 2198 contaminated sites listed (∼32%) without data on contaminants, and in Superfund site data there were 940 sites (∼76%) without contaminant data.

Human biomarkers and wildlife were the most common media tested for contaminants in peer-reviewed literature. While grey literature did not measure or detect contamination, water, food, and sediment were the most common contaminated media mentioned. Both Superfund and FCSI datasets reported contaminant media tested, with soil being the most common in both databases, followed by groundwater (Table 2). There were several media utilized in scientific research that were not identified in federal site assessments. These include human biomarkers (Denham et al., 2005; Fitzgerald et al., 1996; Fitzgerald et al., 2004; Goncharov et al., 2008; Kegler & Malcoe, 2004; Michelle C Kegler et al., 2010; Rock et al., 2019; Tsuji et al., 2005), plants and plant foods (Fitzgerald et al., 1996; Fitzgerald et al., 2004; Garvin, 2018; Koch et al., 2013; Samuel-Nakamura et al., 2017; Sarkar et al., 2019), wildlife and wild game (Brown et al., 2014; Koch et al., 2013; Rock et al., 2019; Samuel-Nakamura et al., 2017), and tree bark (Flett et al., 2021).

**Table 2:** Number of contaminants reported by media type in peer-reviewed articles and federal site records

| Contaminant Media Type | # of contaminants detected per media in peer-reviewed articles<sup>a</sup> (% of total) | # of contaminants reported per media in FCSI data<sup>b</sup> (% of total) | # of contaminants reported per media in Superfund data<sup>b</sup> (% of total) |
| --- | --- | --- | --- |
| Soil (incl. surface soil) | 4 (6.3%) | 3493 (62.8%) | 3817 (34.7%) |
| Groundwater | n/a | 1022 (18.4%) | 2618 (23.8%) |
| Sediment | 3 (4.8%) | 111 (2.0%) | 1841 (16.8%) |
| Surface water | 9 (14.3%) | 133 (2.4%) | 903 (8.2%) |
| Solid Waste | 1 (1.6%) | Not reported | 403 (3.7%) |
| Air | n/a | 56 (1.0%) | 184 (1.7%) |
| Other Media | 46 (73%) | n/a | n/a |
| **TOTAL #** | 63 | 5559 | 10 988 |

<sup>a</sup>To display data that is comparable to federal governmental databases, this column reflects the number of contaminants detected per media per peer-reviewed article collecting primary quantitative data. For a detailed chart on contaminants sampled including the number of samples taken in each study, see supplemental materials (Excel Table S1).
<sup>b</sup>Federal databases (both FCSI and Superfund) provided information on the name of contaminants or contaminant groups found at each site, and the media in which they were found. Notably, the databases do not indicate the number of samples taken per media or the contaminant level. This number therefore provides us with an idea of the prevalence at which each media is sampled, rather than the exact number of samples per media
Note: See Excel Tables S1, S3, and S5 for complete datasets (EPA, 2021d; Goncharov et al., 2008)

### 3.4 Risks posed by contaminated sites

Both qualitative and quantitative data from the three evidence streams provided information on contaminated sites and Indigenous people, land, and food systems. Although a causal link to health outcomes cannot be inferred through federal contaminated site databases, information such as prioritization rankings add to our understanding of the issue’s scope.

#### 3.4.1 Environmental risk assessment

Six peer-reviewed studies focused on the assessment of contaminants found on Indigenous lands affected by contaminated sites (summarized in Table S4), and in five of these studies there was evidence of contaminant levels exceeding regulatory guidelines. A study on an abandoned mine waste site proximate to the lands of the Navajo Nation (Blue Gap Chapter, Arizona), revealed elevated levels of Uranium (67-170 μg/L-1, US EPA max. 30 μg/L-1) in spring water, and detected Uranium (6,614 mg/kg−1), Vanadium (15,814 mg/kg−1), and Arsenic (40 mg/kg−1) in mine waste solids (Blake et al., 2015). Another study examined tree bark on the Spokane reservation (Washington), nearby the Midnight Mine Superfund site, finding a high geo-accumulation index for Uranium and a moderate index for Thorium (Flett et al., 2021). Uranium was also detected in stream sediments on Inuit Territory in Labrador, Canada, proximal to the Abandoned Kitts-U mine site (up to 214.46 mg/kg detected, exceeding the max. of 23 mg/kg) (Sarkar et al., 2019). Sediment toxicity tests in Inukjuak Inuit Territory (Saglek Bay, Labrador) near a military radar site revealed sediment PCB concentrations exceeding Canadian sediment quality guidelines by 41-fold (Brown et al., 2013). Local birds in Saglek Bay, the shorthorn sculpin and black guillemot, were found to be exposed to sediments with concentrations measuring 1000 ng/g within 3 km of a contaminated marine sediment site, exceeding the limits associated with risk to survival (750 ng/g for sculpin and 77 ng/g for guillemot) (Brown et al., 2013). Other studies examined the presence or origin of contaminants. For example, a study on the Yurok Indian Reservation’s Klamath watershed (California) detected a wide variety of contaminants in water, including carbamates, dioxins/furans, mercury, microcystins, organochloride pesticides, and phenols including PCP and TCP (Middleton et al., 2019). The origin of copper contamination dispersed onto the land of the L’anse Indian tribal lands (Michigan) were confirmed to be from the Mass Mill Superfund close to the Keeweenaw Peninsula in one study using sediment core dating (Kerfoot et al., 2020).

#### 3.4.2 Contaminated Sites and Indigenous Food Systems

Seven peer-reviewed articles examined contaminants and Indigenous peoples’ food (both plant and wildlife) (Table S5) (Brown et al., 2014; Fitzgerald et al., 2004; Garvin, 2018; Koch et al., 2013; Rock et al., 2019; Samuel-Nakamura et al., 2017; Schmitt et al., 2006). A total of 58 varieties of plant and animal foods were sampled, including 42 plant species, four mushroom species, and 12 animal species. Seals that are a part of Inuit diets in Labrador, Canada were evidenced to contain PCBs and organochloride pesticides that exceeded adverse effects thresholds of 1.3 mg/kg (Brown et al., 2014). Hares, mushrooms, and wild berries at a variety of contaminated sites affecting First Nations in Canada were assessed, evidencing arsenic bioaccessibility in hare meat and mushrooms (Koch et al., 2013). Mutton consumed by Navajo Nation members in Arizona were found to exceed reference dietary intake levels for uranium, arsenic, cadmium, lead, molybdenum, and selenium (Rock et al., 2019; Samuel-Nakamura et al., 2017). In studies on the traditional foods of the Eight Tribes and Nations of Northeastern Oklahoma, fish and crayfish were found to contain lead and cadmium, posing a hazard to human consumers of carnivorous wildlife (lead consumption from crayfish up to 58.75 mg/kg/day (Toxicological Reference Value (TRV) 1.68), from carp up to 7.54 mg/kg/day; and cadmium from crayfish up to 1.66 mg/kg/day (TRV 1.47) (Schmitt et al., 2006), and 36 species of edible plants within a contaminated area were found to have significantly different levels of cadmium, lead, and zinc than plants outside of the contaminated area (Garvin, 2018). Overall, contaminants studied in food were primarily heavy metals (Garvin, 2018; Koch et al., 2013; Rock et al., 2019; Samuel-Nakamura et al., 2017; Schmitt et al., 2006), with one study examining PCBs and organochloride pesticides (Brown et al., 2014). Traditional foods and medicinal plants are not included as sampling media in Federal site data for Canada or the US (EPA, 2021d; Government of Canada, 2021).

#### 3.4.3 Contaminated Sites and the Health of Indigenous Peoples

Superfund data showed that 59% of contaminants found at Superfund sites were at levels that pose a risk to human health (EPA, 2021c). The Superfund database inventories 122 sites at which human exposures are “under control” (i.e., the exposure is under EPA limits or precautions have been put in place to prevent human exposure), while the human exposure status of 1075 sites remain unknown (EPA, 2021d).

In scholarly literature, a total of 3742 Indigenous participants were studied across articles, with a total of 2224 participants in seven articles related to contaminant exposures and human health outcomes (summarized in Table S6), primarily from the Akwesasne Mohawk (Ontario, Quebec, and New York) (823 participants in 5 studies) and Navajo (Utah, New Mexico, and Arizona) (1304 participants in one study) Nations, and also from the Ramapough Lunaape Nation (New Jersey) (97 participants in one study) (Denham et al., 2005; Fitzgerald et al., 1996; Fitzgerald et al., 2004; Goncharov et al., 2008; Hund et al., 2015; Hwang et al., 2001; Meltzer et al., 2020). The majority of these articles examined PCBs (Denham et al., 2005; Fitzgerald et al., 1996; Fitzgerald et al., 2004; Goncharov et al., 2008; Hwang et al., 2001). A study conducted in the Mohawk community of Akwesasne evidenced the presence of PCBs in breast milk (Hwang et al., 2001) and serum (Fitzgerald et al., 2004) in a sample of 97 Akwesasne Mohawk women.

Among a sample of 138 Akwesasne Mohawk girls, low levels of serum lead (mean 0.49 ug/dL) were associated with significantly lower probability of having reached menarche, and serum PCBs (mean 0.12 ppb) were associated with a significantly higher probability of attaining menarche (Denham et al., 2005). Serum PCBs and pesticides were associated with heart disease in a sample of 335 Akwesasne Mohawk men (Goncharov et al., 2008). A large cohort study (1304 participants) examined chronic diseases in Navajo Nation members, finding self-reported kidney disease, diabetes, and hypertension to be highly prevalent in the population (43% of the sample had at least one of the three diseases, and 20% had at least two), and to be associated with self-reported exposure to Uranium mine wastes (with a 28% higher risk of hypertension for those with an active exposure to mine wastes) (Hund et al., 2015).

Furthermore, four peer-reviewed articles explored the impacts of contaminated sites on Indigenous peoples using qualitative interviews (Table S7) (Cassady, 2007; Hoover, 2013; Smith et al., 2010; Teufel-Shone et al., 2021). In the case of the Aamjiwnaang First Nation (Ontario), semi-structured interviews of 18 community members were conducted, with interviewees reporting fear related to contamination, unknown long-term health effects, and a changing relationship to the earth resulting from a large contaminated site nearby (“Chemical Valley”) (Smith et al., 2010). According to participants, these effects were not addressed by federal cleanup strategies, and continued to impact the community once the site was remediated (Smith et al., 2010). Loss of culture, language, and social connection was reported as an implication of Superfund sites affecting fishing activities in semi-structured interviews with 65 Akwesasne Mohawk community members (Hoover, 2013). Navajo Nation members in 12 at-home focus group interviews described that the Gold King Mine Spill (San Juan River, Northwestern US) was a continuation of colonial violence on Indigenous peoples which removed the community’s agency and voice and forced their relocation (Teufel-Shone et al., 2021). An ethnographic article that involved informal interviews with Inupiak Inuit members (Alaska) described that the discovery of a contaminated site created suspicions about health effects such as cancer amongst the community (Cassady, 2007).

News articles in grey literature provided information on some Indigenous community members’ perceptions of the health impacts of contaminated sites. For example, one article described a shift away from cultural practices due to a large contaminated site (“Chemical Valley”) close to the Aamjiwnaang First Nation (Bienkowski, 2012), and another described the perceived threat of Superfund Sites to the overall health (including both cancer and non-cancer endpoints) of Native American people across the US (Hansen, 2018). A review article in the grey literature also described concerns of Native American communities on the potential effects of abandoned mine sites in the Western United States on child development (Lewis et al., 2015).

### 3.5 Contaminated Sites Management

According to government documents in the grey literature, both US and Canadian contaminated site management programs follow a standard process for assessing, planning for, and remediating contaminated sites (Table S8). Contaminated sites on Indigenous lands are managed similarly to sites on non-Indigenous lands in North America, including the application of standardized risk assessment and management frameworks that fall under federal programs such as the Comprehensive Environmental Response, Compensation, and Liability Act (CERCLA) in the US and Contaminated Sites Management Program (CSMP) in Canada (Contaminated Sites Management Working Group, 2000; EPA, 2021c). These programs provide guidelines for site classification and prioritization and outline a step-wise process for addressing federal contaminated sites.

Under these programs, 37.5% of Superfund sites are classified as “active” while the remainder have been “archived” in the federal data. Similarly, 35.3% of Canadian sites are categorized as “active” while 23.4% are “suspected” and the remainder are “closed”. FCSI data indicated the current stage of contaminated site management (Figure 4), with the largest proportion of active sites (n=1331) in the “detailed testing” stage. Superfund data reported on whether the site required “no further action/not eligible” (688 records), “referred to a cleanup program” (277 sites), or “assessment needed or ongoing” (62 sites). While it was not possible to categorize all 49 peer-reviewed articles by stages of contaminated site processes, these were tagged by the author based on the topic of focus, which was most commonly risk assessment (11 articles), environmental assessment (9 articles), risk management (9 articles), and socio-cultural impacts (8 articles), with fewer articles focused on post-remediation (3 articles) and long-term assessment/legacy impacts (1 article).

**Figure 4:**
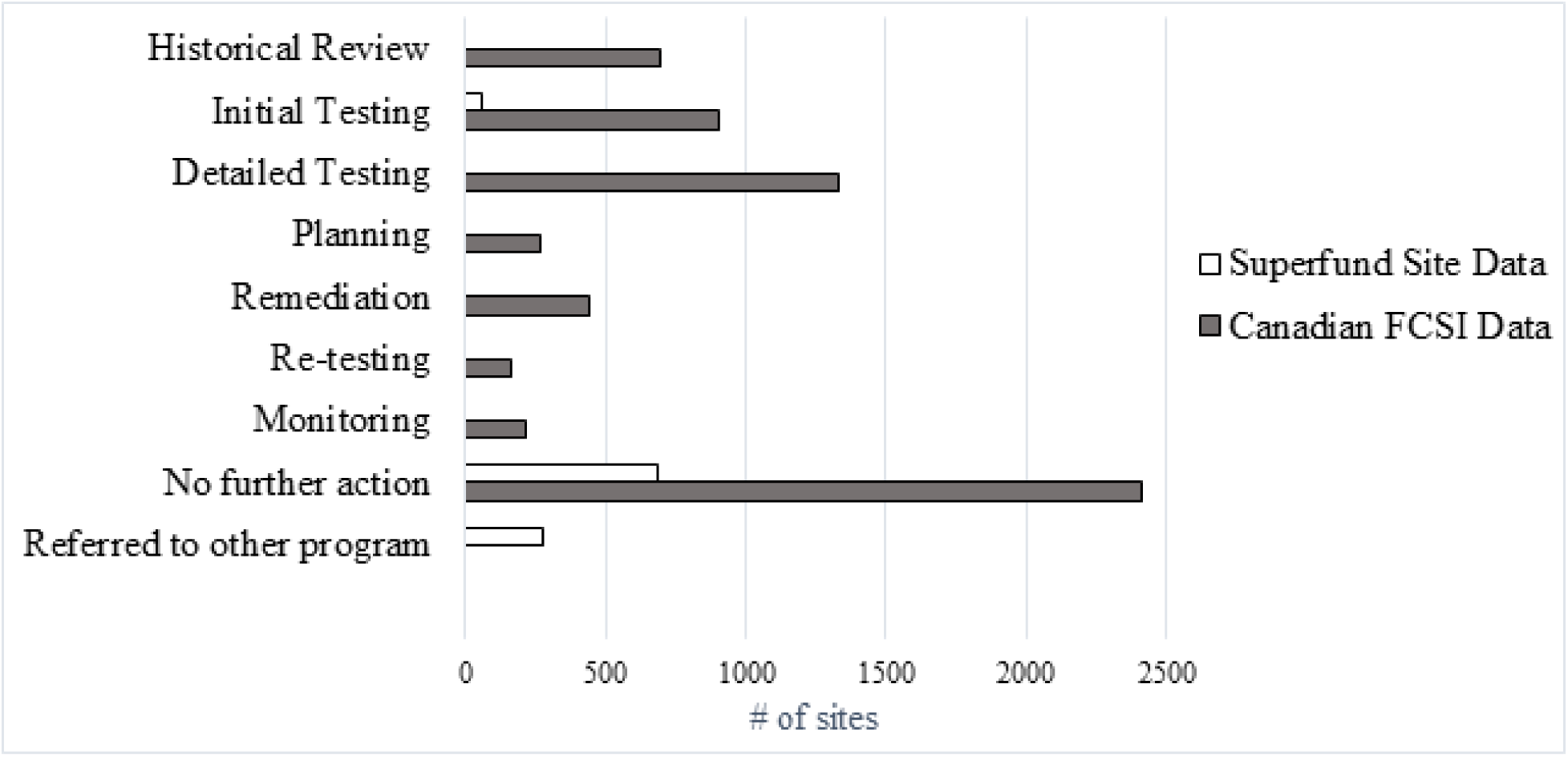
Bar graph representing the number of sites at each stage of the contaminated sites management process for Canada and US Data, classified under federal management programs (CERCLA and CSMP) (Excel Tables S3 and S4)

### 3.6 Health Risk and Exposure Assessment

In an effort to address the unique consumption patterns of subsistence populations, federal governments have published guidelines for more comprehensively assessing exposures to country foods, including in the context of frameworks such as the Human Health Risk Assessment (HHRA) and through the development of exposure scenarios (Harper, 2007; Health Canada, 2018). In peer-reviewed literature, the benefits and challenges of assessments for contaminated sites that include exposure scenarios for Indigenous communities were outlined in two primary and four secondary articles (Tables S9 and S10). Potential benefits were described in two case studies on the testing and validation of exposure scenarios related to hazardous waste contamination on the Umatilla Indian reservation (Oregon), and of uranium contamination on the Spokane reservation (Washington). These exposure scenarios were found to improve the holistic nature of risk assessments, which included the consideration of pre-site land use and socio-political factors associated with exposure, compared to generic risk assessments that do not measure community-specific exposure (Harper et al., 2012; Harper et al., 2002). Challenges to risk assessment were described in two peer-reviewed articles on the development of exposure scenarios for consumption patterns of the Confederated Tribes of the Umatilla Indian Reservation (Oregon) located near a Superfund (Department of Energy’s Hanford Site) (Harris & Harper, 1997), and the validation of a risk assessment ingestion value by estimating soil ingestion by the Xeni Gwet’in First Nation (British Colombia) through the contaminated Chilko Watershed (Doyle et al., 2012). Both studies found inconsistencies between risks posed to Indigenous peoples and risks assessed in standard HHRAs, including soil ingestion values and exposures due to lifestyle factors, religious practices, and non-food exposures (Doyle et al., 2012; Harris & Harper, 1997). In qualitative research, interviews with 35 members of the Colombia river basin tribe (Umatilla, Oregon) highlighted that risk assessments in Indigenous communities measure risk on the same scales as non-Indigenous communities, despite evidence that contaminated sites create unique risks for Indigenous people that are not possible to quantify using traditional methods (Harris & Harper, 1997). Furthermore, two discussion articles critiqued standardized risk assessments due to a lack of consideration of cumulative effects of contaminants, incompatibility with tribal life ways (Holifield, 2012), the privileging of property owners and lack of benefit to Indigenous peoples, and the use of non-Indigenous definitions of health to define risks (Arquette et al., 2002). An article focused in the Akwesasne Mohawk community cited a lack of Indigenous involvement and resources allocated to the community as a major barrier to effective risk assessment (Arquette et al., 2002).

Grey literature evidenced risk assessments that were conducted for specific sites and communities, including for example an HHRA of legacy arsenic contamination as a response to concerns surrounding health risks from the Ndilo and Dettah (Dene) communities (Yellowknife, Northwest Territories).(Government of Northwest Territories) According to government documents, under CERCLA and CSMP, health risk assessment is used to measure and quantify physical health risks and is part of the standardized process for contaminated site management. Notably, according to Canadian guidelines, conducting an HHRA typically requires the “hire of a qualified consultant with the necessary technical and scientific expertise to perform the work” (Contaminated Sites Management Working Group, 2000). In Canada, the National Classification System for Contaminated Sites is used to evaluate and classify sites based on the level of human and environmental risk posed by the site (Canadian Council of Ministers of the Environment, 2008). In Canadian FCSI Data, sites are ranked as either “High priority” (1016 site records), “Medium priority” (1198 site records), “Low priority” (491 site records), or “No priority for action” (429 site records) based on this system (Government of Canada, 2021). A site classified as “High priority” has imminent risks or documented adverse outcomes to both human and environmental health (Canadian Council of Ministers of the Environment, 2008). The US EPA uses a similar tool, the Hazard Ranking System, to assign a quantitative value to the human and environmental risks posed by a site, which determines whether a Superfund site will be categorized under a “National Priorities List” (EPA, 2017). The Superfund dataset categorized site records as “not NPL” (1022 records), “deleted NPL” (27 records), “removed from NPL” (one record), “proposed NPL” (nine records), “part of an NPL site” (46 records), or “final NPL” (131), indicating that 186 sites are of high priority (i.e., classified as “NPL) (EPA, 2021d). Thus, across Canadian and US federal datasets, there are 1202 sites that are classified as “high priority” due to human and environmental health risks (Chan, Fediuk, et al., 2021; Samuel-Nakamura et al., 2017).

### 3.7 Risk Management

The management of risks related to contaminated sites in Canada and the US involves removing or reducing contaminants and limiting the use of contaminated media by affected communities. Risk management strategies are unique to each site, and involve input from regulators, site owners, policies, and local communities (Contaminated Sites Management Working Group, 2000; EPA, 2022d). Five peer-reviewed research articles tested various risk management strategies related to contaminated sites and Indigenous communities (Table S11). A study on risk mapping of contaminants on the lands of the Navajo Nation (Churchrock Chapter, New Mexico) and a survey of 151 community members found that the maps were helpful in avoiding the risk of consumption of contaminated food and water arising from the Churchrock Uranium Mine Site (deLemos et al., 2009). Another study cited the effectiveness of safety protocols (education on the proper handling of hazardous wastes) for nine Fort Albany Cree First Nation workers (Ontario) employed to clean-up the Mid-Canada Radar Contaminated site, finding no significant burden of blood lead and PCBs before and after the clean-up (Tsuji et al., 2005). Three studies explored the influence of a lay-health advisor intervention on blood lead concentrations in children living in proximity to the Tar Creek Superfund Site, comparing results between members of the Eight Tribes and Nations of Northeastern Oklahoma and Non-Indigenous, white participants that lived near the site. Influences on lead exposure prevention behaviors were first assessed through qualitative interviews with 380 children’s caregivers (Bland et al., 2005). Then, caregiver-child pairs (n=331 pairs, 43.5% Native American) participated in a cross-sectional study, measuring children’s blood lead, and conducting structured caregiver interviews before and after a 2-year intervention in which caregivers were trained by local community health advisors on how to prevent children from blood lead poisoning (Bland et al., 2005; Kegler & Malcoe, 2004). The study found a reduction in blood lead before and after the intervention. Of Native American children, blood lead levels decreased significantly from time 1 (6.00 ug/dL) to time 2 (4.97 ug/dL), with no significant difference from the white control group. Another study was conducted at a third time point (4 years later), finding improved lead preventative behaviors (handwashing) in Indigenous participants (167 Indigenous and 213 white participants) (Michelle C Kegler et al., 2010). Another review article described a precautionary principle for risk management at the Zortman-Landusky Mine Superfund Site on the Fort Belknap Reservation (Montana), home to the Assiniboine and Gros Ventre Tribes, in which caution was exercised in site management when risk is unknown (Emel & Krueger, 2003).

Five peer-reviewed articles described the use of consumption advisories to manage risks related to country foods. One article focused on the territories of the Ahahminquus community of the Mowachaht Tribe and the Silammon people (British Colombia) and analyzed the use of Canadian health advisories and fishery closures related to contaminated sites near the Gold river and Powell river fisheries. The study found that the risks are comparable when switching from country foods to market foods, and that such advisories may be substituting one risk for another (Wiseman & Gobas, 2002). The relationship between country foods and market foods was also examined in two studies focused on the Inupiaq Inuit and the Akwesasne Mohawk Nation (Cassady, 2007; Hoover, 2013). Advisories for consuming traditional fish resulted in inadequate nutrition and cultural loss for the Akwesasne Mohawk Community (Hoover, 2013). An ethnographic field study focused on the Project Chariot abandoned waste site (Alaska) described that Inupiaq Inuit considered traditional foods to be curative and preventive of health issues, despite the issuance of advisories, and also described local knowledge (e.g., visually checking wild game for tumors) that could be used to determine if food was contaminated (Cassady, 2007). Two articles in grey literature discussed the contamination of subsistence food resources in Indigenous communities in general (Bienkowski, 2012; EPA, 2015), with one article noting that studies of contaminants most often result in avoidance advisories (Bienkowski, 2012). One government document described successful implementations of consumption advisories and community education related to contaminated foods on the lands of several Indigenous communities after receiving federal grants from the US EPA Superfund, including the Akwesasne Mohawk Nation, Anishinaabe Nation (Great lakes region of Canada and the US), and Yurok tribe (California) (EPA, 2015).

### 3.8 Long-Term Management

In peer-reviewed literature, information in five review articles described that communities continue to face long-term site-related challenges after a site is remediated and closed by the federal government (Moore-Nall, 2015; J. Sandlos & A. Keeling, 2016; John Sandlos & Arn Keeling, 2016; Smith et al., 2010; Teufel-Shone et al., 2021). For example, one article described that a contaminated site resulted in alienation of Navajo Nation members (Arizona) from their traditional territory (John Sandlos & Arn Keeling, 2016). Another article described that there was a lack of acknowledgement of the legacy impacts of contaminated sites by federal governments, despite advocacy efforts by the Dettah and Ndilo communities (Yellowknife) on the remediation of the Giant Mine in Canada (J. Sandlos & A. Keeling, 2016). Long-term health impacts were also described in the context of the Los Alamos National Laboratory Superfund Site and the Tribes of the Southwestern US (Moore-Nall, 2015). The Gold King Mine spill was described in one review article as contributing to a history of relocation and lack of agency and voice for the Navajo (Dine) Nation, with lasting impacts on the community (Teufel-Shone et al., 2021).

Three semi-quantitative studies examined post-remediation land uses of contaminated sites (Table S12) (Burger, 2004a, 2004b; LeClerc & Keeling, 2015). Two of these studies utilized structured interviews and found that future land use preferences differed by ethnicity, with participants that were members of the Shoshone Bannock tribe (Idaho) rating camping, fishing, hunting, and returning the land to Native people higher than white participants did. Native American participants were generally underrepresented, however, in both studies (324 out of 1370 participants (23.6%) and 11 out of 254 participants (3.5%)) (Burger, 2004a, 2004b). Another study involving semi-structured interviews of 18 members of the Dene and Metis communities of Fort Resolution (Northwest Territories) found that since the establishment and closing of the Abandoned Pine Point Mine site, the community’s land use patterns had changed from a land-based economy to a mixed economy reliant on wage labor, concluding that post-remediation land use would likely be different than land uses before the site’s establishment (LeClerc & Keeling, 2015).

In conference materials in grey literature, one document recommended designing remediation processes at Superfund sites that can more effectively account for tribal needs to permanently restore the land for tribal uses (Michelsen, 2010). Another article described that due to the political and financial dynamics associated with site remediation on reserve land in Canada, the federal government will remediate a site at the lowest possible cost, which often does not meet the standard that Indigenous communities desire for post-remediation land use and development (Gailus, 2013). In federal datasets, both Superfund and FCSI categorized most of the sites under “no further action required”, with 2408 such sites in Canada and 688 sites in the US (EPA, 2021d; Government of Canada, 2021).

### 3.9 Indigenous Inclusion, Collaboration, and Leadership

#### 3.9.1 Collaborative research

Of the included peer-reviewed literature, 20 articles described a collaboration between academic researchers and Indigenous people or communities (Arquette et al., 2002; Blake et al., 2015; Denham et al., 2005; Fitzgerald et al., 1996; Fitzgerald et al., 2004; Flett et al., 2021; Goncharov et al., 2008; Harper et al., 2012; Hoover, 2013, 2016; Kegler & Malcoe, 2004; Michelle C Kegler et al., 2010; Michelle C. Kegler et al., 2010; Meltzer et al., 2020; Middleton et al., 2019; Rock et al., 2019; Sarkar et al., 2019; Smith et al., 2010; Teufel-Shone et al., 2021). Several articles described collaborative efforts between academic institutions and Indigenous communities, as well as community-led research projects (deLemos et al., 2009; Goncharov et al., 2008; Harper et al., 2012; Hoover, 2013, 2016; Kegler & Malcoe, 2004; Michelle C Kegler et al., 2010; Michelle C. Kegler et al., 2010; Rock et al., 2019). Some of these studies described such collaborations as mutually beneficial. For example, interviews with 64 Akwesasne Mohawk members on their collaboration with SUNY Albany, a public University in New York State, revealed that there were benefits to members such as education, job skills, grant money and information; and to researchers such as better results, access to the community and help of Mohawk workers in the field (Hoover, 2016). One article analyzed contaminated site response networks consisting of the Eight tribes of Northeastern Oklahoma and non-Indigenous organizations, concluding that there was an increase in collaboration over time (Michelle C. Kegler et al., 2010). Peer-reviewed literature supports that federal grants help to facilitate multi-study projects in communities, as was the case with a collaborative project between the Eight tribes of Northeastern Oklahoma and the University of Oklahoma entitled ‘Tribal Efforts Against Lead’ (Kegler & Malcoe, 2004; Michelle C Kegler et al., 2010; Michelle C. Kegler et al., 2010). Challenges to collaboration were also described in peer-reviewed literature. For example, members of the Akwesasne Mohawk community described difficulties with time constraints due to the finite nature of academic funding, a lack of trust between researchers and community members, and inadequate science communication (Hoover, 2016). Two review articles described an ongoing need for greater funding and resources allocated to collaborative Indigenous-lead research on contaminated sites in the United States (Lewis et al., 2017; Moore-Nall, 2015).

Grey literature and government websites indicated that federal funding agencies in both the US and Canada provide support to Indigenous communities to address contaminated sites (EPA, 2022c; 2021), which has helped to build capacity and resilience (Environment and Climate Change Canada, 2019; Hoover, 2016; United States Government Accountability Office, 2020). Examples of such programs include the First Nations Environmental Contaminants Program (FNECP) and the Northern Contaminants Program (NCP) in Canada, and funds allocated under CERCLA in the US, including the Superfund State and Indian Tribe Core Program Cooperative Agreements (EPA, 2022b). Government reports in grey literature describe that tribes in the US have used EPA grants to support capacity building in environmental programs, and that First Nations in Canada have benefitted from the FNECP (Environment and Climate Change Canada, 2019; Hoover, 2016; United States Government Accountability Office, 2020). Many federal granting agencies require that Indigenous communities partner with academic institutions (Ferguson, 2021; Indigenous Services Canada, 2021; National Institute of Environmental Health Sciences, 2022), which is aligned with the aforementioned peer-reviewed literature from academics involving collaborative efforts with Indigenous communities.

Furthermore, grey literature from governmental funding agencies and review papers described that the requirement to partner with an established scientist trained in an academic institution (Ferguson, 2021; Indigenous Services Canada, 2021; National Institute of Environmental Health Sciences, 2022) limits the ability of many communities to obtain funding independently, and often results in a large amount of scientific and monetary resources focused on a single community (Ferguson, 2021; Fitzgerald et al., 1996; Fitzgerald et al., 2004; Hoover, 2013, 2016; Kegler & Malcoe, 2004; Michelle C Kegler et al., 2010), while other sites have minimal support.

#### 3.9.2 Collaborative Site Management

Peer-reviewed literature described the methods of evaluation collaborations between Indigenous communities and non-Indigenous institutions, such as in the example of the Fort Albany Cree First Nation, wherein a Canadian governmental framework was used to evaluate whether a true partnership existed between the Canadian government and First Nation (Sistili et al., 2006), Another review article described a diminished societal, governmental, academic, and political response to the Sequoyah Corporation fuels release and the Church Rock spill, nuclear releases affecting the Cherokee (Oklahoma) and Navajo (New Mexico) nations, compared to contaminating events of a similar scale affecting white communities (Brugge et al., 2007).

Government documents are available in grey literature on inclusion and collaboration with Indigenous communities on contaminated site processes. Canada has a federal guidance document indicating areas in which inclusion of Indigenous communities may occur, recommending that Indigenous people assist with the contaminated site process, and providing potential opportunities for Indigenous involvement (for example, community members performing media sampling or hiring Indigenous companies to carry out remediation work) (Health Canada, 2010). Indigenous inclusion on Superfund site management follows a consultation process outlined by the US EPA, which defines the establishment, appropriateness and extent of consultation and collaboration with federally recognized tribes.(EPA, 2011a) After determining that there was insufficient consultation with tribes at NPL Superfund sites (18 NPL sites with documented consultation, 7 of which had incomplete data), the US Government Accountability Office recommended documented consultation with tribes at 4 of the 9 steps in contaminated site processes, which excludes initial assessment, remedial design, construction completion and post-construction completion (i.e., maintenance and long term actions) and post-remediation development (US Government Accountability Office, 2019).

Non-governmental grey literature highlighted a lack of Indigenous inclusion in contaminated sites assessment and management processes voiced by Indigenous peoples. For example, a news article described that members of the Akwesasne Mohawk Tribe (New York) were dissatisfied with their opportunities to provide input into remediation decisions on the US EPA Alcoa Grasse River Superfund site (Indian Country Today, 2013). Chief Glenn Nolan of the Missanabie Cree First Nation (Ontario) outlined the lack of Indigenous involvement in the assessment, management, and communication of risks associated with abandoned mine sites (Nolan, 2009). The article proposed actions to improve consultation with Indigenous people, including community involvement beginning from initial assessment, accessible communication of results, and the development of ongoing collaborative monitoring strategies (Nolan, 2009).

While not a dominant theme in literature or government guidance documents, successful instances of Indigenous leadership in contaminated site management are evidenced in grey literature sources (Assembly of First Nations, 2001; EPA, 1996; Tribal Superfund Working Group, 2022). The Tribal Superfund Working Group, for example, has published several tribe-led efforts to fight community contamination, such as in the case of the Fort Mojave Indian Tribe (California) remediation project on the Topock site (Tribal Superfund Working Group, 2022). First Nations in Canada have also demonstrated leadership in reclaiming and remediating land contaminated by abandoned mines, for example, by leading discussions among the Assembly of First Nations (Canada-wide) on remediation options (Assembly of First Nations, 2001).

#### 3.9.3 Traditional Knowledge and Contaminated Sites

In peer-reviewed literature, one discussion article described the negative impacts of historical subsumption of Traditional Knowledge within technical contaminated site processes in the remediation of the Giant Mine, Northwest Territories, which was seen by the local Dene First Nation, Dettah and Ndilo (Navajo) communities as an inadequate and inappropriate inclusion of Traditional Knowledge (J. Sandlos & A. Keeling, 2016). The US EPA recently published a document outlining the ‘integration’ of Traditional Knowledge into environmental science, policy, and decision-making processes, in response to tribal leaders’ requests to enhance its use (Woolford, 2017). In grey literature, Health Canada lists Traditional Knowledge as a component of exposure assessment within the HHRA (Health Canada, 2018), and in a recent update of their Northern Contaminated Sites Management Plan (NCSMP), pledged to increase inclusion of Traditional Knowledge studies into project planning and implementation, although the operational methods of this are not published (Crown-Indigenous and Northern Affairs Canada, 2021).

## 4. DISCUSSION

This scoping review identified, mapped, and analyzed the existing information on contaminated sites and Indigenous peoples in Canada and the US, and sought to address the three main objectives outlined in the introduction. The research was motivated by observations that contaminated sites disproportionately affect Indigenous communities (Fernández-Llamazares et al., 2020; Lewis et al., 2017), and that there is limited research on the subject matter, without which a deeper understanding cannot be realized to permit evidence-based solutions. Findings from 49 scholarly articles, 20 grey literature pieces, and US and Canadian federal contaminated site databases revealed a disparate yet also vast and multi-disciplinary body of information on contaminated sites affecting Indigenous peoples and their health, land, and food systems. The number of articles identified from the systematic literature search covers a relatively small proportion of the total number of federal contaminated sites identified from federal inventories that affect Indigenous peoples. Of the peer-reviewed literature retrieved, a total of 35 sites were described, while there were 8114 sites inventoried by federal governments. Information on 815 Indigenous communities was identified through this review, while there exist in total 574 Tribal Entities in the US (National Conference of State Legislators, 2020) and in Canada there are 630 First Nations and 53 Inuit communities (Crown-Indigenous and Northern Affairs Canada, 2022), as well as eight Metis Settlements (although the majority of Metis in Canada do not live on official settlements). Thus, the information available identifies a substantial amount (approximately 64%) of Indigenous communities that are potentially impacted by federal contaminated sites, though this is likely an underestimation.

We note through our mapping exercise that site locations in the US and Canada are widespread, however the majority are south of the 60^th^ parallel in Canada, and that data from the three evidence streams may largely underrepresent the geographic scope of contaminated sites that affect Indigenous peoples. A discussion specific to federal contaminated sites is applicable to Indigenous communities that live on federally managed land. Notably, due to regulatory and legal provisions under the Indian Act, Canadian FCSI data primarily concerns First Nations reserves, which are considered federal lands and are mostly inhabited by First Nations communities with the majority being south of the 60^th^ parallel. Approximately 328,048 (40 %) of First Nations members in Canada live on reserve land according to the 2016 census (Indigenous Services Canada, 2020). Accordingly, FCSI data does not represent the total amount of contaminated sites affecting Indigenous communities in the country, and furthermore, Inuit and Metis communities are likely most underrepresented, as the majority do not live on reserve land (Gailus, 2013). Contaminated sites affecting Indigenous communities off-reserve are usually managed provincially or privately and thus are not captured by federal databases (Government of Canada, 1985). One grey literature article estimated that 1200 Indigenous communities in Canada have either an active mine, an abandoned mine site, or a mine exploration project on their territory, which is more than double the number of communities listed in the FCSI, demonstrating that there are many sites of potential or actual concern to Indigenous communities that are not tracked by the federal government (Nolan, 2009). Similarly, the US EPA does not have explicit criteria for determining how a Superfund Site is deemed to be of ‘Native American Interest’, and many sites are managed by non-federal governing bodies (termed ‘cleanups’) or private industries (EPA, 2021b; US Government Accountability Office, 2019), which are not included in Superfund data. Thus, the information presented in this review is limited to federally managed and inventoried contaminated sites affecting Indigenous peoples, but there are many sites managed under other jurisdictions not accounted for by federal governments or this review.

Contaminated sites are a pervasive issue for Indigenous communities in Canada and the US, with peer-reviewed literature and federal data indicating that the largest proportion of these sites arise from mining activity and industrial development on or adjacent to lands inhabited by Indigenous peoples. It is notable that there are many sites for which the source of contamination is unknown or not documented, for example, 248 (20%) of Superfund site records do not inventory the source of contamination, and FCSI does not inventory contaminant sources. This review also found that there is minimal primary scientific research on the assessment of contamination from these sites (i.e., 45 unique contaminants measured in scientific studies in the literature compared to 440 unique contaminants measured and recorded in federal datasets), with most studies examining a single contaminant or a handful of contaminants.

Indigenous peoples are exposed to contaminants from contaminated sites through a variety of pathways, which includes inhalation, ingestion of drinking water and contaminated foods, and contact with contaminated soil and sediments. Literature on traditional food systems across Canada and the US evidences a diversity of exposure pathways unique to Indigenous communities, including for example through subsistence diets that include wild foods and medicinal plants (Chan, Fediuk, et al., 2021; Fernández-Llamazares et al., 2020; Jonasson et al., 2019). Notably, while peer-reviewed literature examined in this review documents the presence of a variety of contaminants in plant foods, wildlife, wild game, and human biomarkers (Brown et al., 2014; Denham et al., 2005; Fitzgerald et al., 1996; Fitzgerald et al., 2004; Flett et al., 2021; Garvin, 2018; Goncharov et al., 2008; Kegler & Malcoe, 2004; Michelle C Kegler et al., 2010; Koch et al., 2013; Rock et al., 2019; Samuel-Nakamura et al., 2017; Sarkar et al., 2019; Tsuji et al., 2005), information on these potential exposure sources is not captured in federal databases.

As Indigenous peoples have unique consumption and land use patterns compared to the general population, exposures that are estimated based on generic federal frameworks employed in environmental assessments may be limited (Arquette et al., 2002; Cassady, 2007; Doyle et al., 2012; Harris & Harper, 1997; Holifield, 2012; Lewis et al., 2017). In a search of the US EPA Exposure Factors Handbook with the keyword “Native American”, six tables are available pertaining to exposure to contaminants through the consumption of fish, but there is no information pertaining to any other exposure pathway (EPA, 2022a). This is one example of the paucity of data and classification tools for environmental assessments involving Indigenous peoples.

Our review found that the current body of literature on the potential health impacts of contaminated sites on Indigenous peoples is relatively small and lacks diversity. We identified seven peer-reviewed articles with quantitative data on health outcomes. Four of these articles were focused on the Mohawk community of Akwesasne (Table S6), and they documented a range of chronic and developmental outcomes associated with exposure to chemical stressors (primarily lead and PCBs), including early menarche, heart disease, kidney disease, diabetes, and hypertension. In four qualitative peer-reviewed studies, culture, spirituality, language, and community were found to be adversely influenced by contaminated site exposure (Cassady, 2007; Hoover, 2013; Smith et al., 2010; Teufel-Shone et al., 2021), and these are social and environmental determinants of Indigenous Peoples’ health (Reading & Wien, 2009). There was evidence that many Indigenous communities face long-term site-related challenges including a changing relationship to the earth, fear related to contamination, and cultural loss (Moore-Nall, 2015; John Sandlos & Arn Keeling, 2016; Smith et al., 2010; Teufel-Shone et al., 2021). While these findings are not easily generalizable (i.e., they report on findings from 6 of 815 communities identified by federal databases who are potentially exposed to contaminated sites), they alert us to the possibility that health risks are elevated in such impacted communities and call for the need to scale-up research in this area.

According to federal data, the progression through the ten-step process for addressing a contaminated site can take decades (EPA, 2021a; Government of Canada, 2021). In the Canadian commissioner’s report on federal contaminated sites, it was identified that there are more sites in the inventory than funds available for their management, and as a result only the highest priority sites are addressed (Ellison, 2012; Government of Canada, 2012). Furthermore, existing regulations make it difficult for communities to receive federal funding for site remediation unless they can prove that there is a high risk to human health and the environment, the assessment of which involves lengthy and expensive technical processes that are often outsourced to experts in academic and consulting, for example, that conduct HHRAs (Gailus, 2013). This results in a catch-22 in which the current regulatory and federal funding structure requires that Indigenous communities rely on outsourcing work, which thus limits internal capacity building.

This review found that contaminated sites are often addressed by federal governments, industry, and the scientific community as an isolated, technical problem, (Contaminated Sites Management Working Group, 2000; EPA, 2011b; 2019) while they are considered by many Indigenous peoples as part of a broader legacy of environmental injustice (Cassady, 2007; Holifield, 2012; Lewis et al., 2017; Moore-Nall, 2015; John Sandlos & Arn Keeling, 2016; Smith et al., 2010; Teufel-Shone et al., 2021). Federal risk assessment systems are useful to organize and prioritize site management, however according to our review these assessments are not necessarily aligned with the needs of the communities affected by them (Contaminated Sites Management Working Group, 2000). Furthermore, there is evidence in peer-reviewed literature to suggest that the use of generic “one-size-fits-all” risk assessment models (such as the HHRA in Canada and the Hazard Ranking System in the US) for contaminated sites lack benefit to Indigenous people (Arquette et al., 2002; Cassady, 2007; Doyle et al., 2012; Harris & Harper, 1997; Holifield, 2012; Lewis et al., 2017). Although sites are ‘closed’ when they are deemed to be at a safe risk level by federal governments, this review found evidence that Indigenous peoples face long-term and cumulative challenges that are not supported by governmental resources and funding (Moore-Nall, 2015; J. Sandlos & A. Keeling, 2016; John Sandlos & Arn Keeling, 2016; Smith et al., 2010; Teufel-Shone et al., 2021). This current western scientific approach contradicts Indigenous epistemologies such as the Seven Generations Principle, a Haudenosaunee philosophy followed by many Indigenous communities in Canada and the US, in which decision-making considers the well-being of people and environments seven generations into the future (Joseph, 2020). These factors point to the need for a holistic approach from both academics and governments which includes moving beyond physical and finite impacts to understanding contaminated sites as complex, multi-faceted issues with long term and cumulative effects (Brugge et al., 2007; Holifield, 2012; Nolan, 2009; J. Sandlos & A. Keeling, 2016; Smith et al., 2010; Teufel-Shone et al., 2021). Recent evidence affirms the positive benefit of environmental risk assessments that privilege Indigenous epistemologies and leadership, a strategy which may help to respond to the lack of prioritization of Indigenous conceptualizations of risk in contaminated site assessment (Buell et al., 2020). Taken together, these factors point to a need for efficient, low-cost, Indigenous-led and community-based assessment methods that can rapidly predict health and environmental risks, which would help to build community capacity and improve the efficiency and cost-effectiveness of current assessment and prioritization processes. There is an increasing focus on the development of New Approach Methodologies (NAMs) for environmental risk assessment by industry, government, and academics (Government of Canada, 2017; Krewski et al., 2010), and these findings suggest that Indigenous perspectives should be considered and prioritized as a part of current NAMs development.

The majority of peer-reviewed research on risk management included in this review was focused on avoiding contaminant exposure (for example, through the use of fish consumption advisories), which is consistent with a global pollution management trend in which risk reduction through public and community level intervention is replaced with risk avoidance that emphasizes individual level action (Fernández-Llamazares et al., 2020). It is possible that this ‘downstream’ approach to management of contaminated sites perpetuates the vulnerability narrative that is often associated with Indigenous communities by Western knowledge systems, having the potential to hinder communities from gaining greater autonomy (Haalboom & Natcher, 2012). However, we note that there are cases where contaminants pose high risks to health and risk avoidance strategies are necessary in the short-term, and in these situations, community involvement may help to improve site management. A recent review indicated that through the involvement of First Nations communities in the process, risk management approaches such as consumption advisories can provide greater benefit to First Nations (McAuley & Knopper, 2011). This is one strategy that would increase the effectiveness of federal contaminated site risk management, and the operationalization of this involvement is an area for further research. We also note that a large proportion of peer-reviewed articles (n=20) identified in this review were problem-based (Blake et al., 2015; Brown et al., 2014; Brown et al., 2013; Cassady, 2007; deLemos et al., 2009; Denham et al., 2005; Fitzgerald et al., 1996; Fitzgerald et al., 2004; Goncharov et al., 2008; Hoover, 2013; Hund et al., 2015; Hwang et al., 2001; Kerfoot et al., 2020; Koch et al., 2013; Meltzer et al., 2020; Middleton et al., 2019; Rock et al., 2019; Samuel-Nakamura et al., 2017; Smith et al., 2010; Teufel-Shone et al., 2021) as opposed to solution-focused. Notably, we did not find any peer-reviewed articles focused on specific remediation strategies nor restoring land back to pre-contamination conditions.

Most research we found on contaminated sites and Indigenous peoples involved a collaboration between western academics and Indigenous communities (Excel Table S1). Several such partnerships, such as in the case of the Mohawk community of Akwesasne and SUNY Albany, have shown to provide mutual benefit (Hoover, 2016). While our review notes that industry, government, and the scientific community acknowledge the importance of Indigenous inclusion (Arquette et al., 2002; Ellison, 2012; Gover, 2007; Holifield, 2012; Michelsen, 2010; Nolan, 2009; Sistili et al., 2006), we found minimal evidence of Indigenous leadership in the management of contaminated sites. Recent literature from Indigenous scholars on natural resource management describes that the inclusion of and collaboration with Indigenous peoples in environmental management is often tokenistic (Parsons et al., 2021). The contribution of Indigenous voices in contaminated site management often occurs too late in the federal process, which perpetuates the “downstream” approach to site management discussed earlier. Vague definitions of ‘inclusion’ of Indigenous peoples in contaminated site management were found throughout this review (Health Canada, 2010), (EPA, 2011a), which risks the subsumption of Indigenous peoples into non-Indigenous management bodies.(Fernández-Llamazares et al., 2020) Our review found that non-Indigenous stakeholders are the powerholders in the management of contaminated sites and largely control collaborations with Indigenous communities. For example, US and Canadian federal guidance documents outline how, when, and to what extent Indigenous peoples are to be included or consulted, omitting Indigenous leadership in designing the process or management plan, and thus maintaining federal governments as the power-holders (EPA, 2011a; Health Canada, 2010). In the US, under CERCLA, tribes are excluded from identifying contaminated sites for inclusion on the Superfund National Priorities List (Gover, 2007). Furthermore, based on our findings, the information related to contaminated site management on Indigenous lands was dominated by non-Indigenous voices. Government documents and academic articles included in this review were primarily authored by non-Indigenous persons, while Indigenous voices could be found in news articles and other web-based grey literature. To this effect, platforms that provide a seat at the decision-making table for federal contaminated sites may underrepresent Indigenous peoples. These findings are consistent with a recent review highlighting the inadequate inclusion of Indigenous peoples in environmental management in general (Fernández-Llamazares et al., 2020). Increased efforts and research to strategize Indigenous leadership and prioritize Indigenous epistemologies in relation to contaminated site management may contribute to more holistic and sustainable contaminated site management processes.

Results indicated that federal governments, Indigenous communities, and academics acknowledge a complex relationship between Indigenous Traditional Knowledge and western science in the context of contaminated site management, but that the approaches to achieving successful collaboration are a work in progress (EPA, 2011a; Health Canada, 2010; Reid et al., 2021; J. Sandlos & A. Keeling, 2016; Woolford, 2017). Recent publications demonstrate that increased efforts are being made to collaborate with Traditional Knowledge holders, including through increased federal funding for collaborations, working groups, and discussions around Traditional Knowledge (EPA, 2011a; J. Sandlos & A. Keeling, 2016; Woolford, 2017). However, federal and academic collaboration initiatives in contaminated site management and research processes may in some cases continue to perpetuate paternalistic environmental practices if left unchecked, as we note that “Using”, “Incorporating”, and “Integrating” Traditional Knowledge into western science and governance has been described in academic literature as a form of assimilation (Reid et al., 2021). New approaches that engage Indigenous and Western ways of knowing harmoniously may continue to propel collaborative engagement between knowledge systems forward. One such approach is Two-Eyed Seeing, a term popularized by Mi’kmaw Elder Dr. Albert Marshall, through which we “learn to see from one eye with the strengths of Indigenous knowledges and ways of knowing, and from the other eye with the strengths of mainstream knowledge, and we use both eyes together” (Reid et al., 2021). Through this approach, Indigenous environmental management goals related to contaminated sites may begin to be realized through collaboration between networks such as Indigenous leadership, Indigenous community members, industry, federal governments, and the scientific community.

### 4.1 Strengths and Limitations

This is the first scoping review to examine the issue of federal contaminated sites and Indigenous peoples, and thus addresses a research gap. Given the disparate evidence base, the compilation and synthesis of data and information from the scholarly literature, federal data and grey literature is a major strength of this review, as it enables us to see with greater diversity, clarity, and objectivity the many perspectives involved in the subject matter. The use of both published and unpublished literature promotes the consideration of voices that are not typically heard in academic literature. Furthermore, the study used a rigorous and transparent search strategy, following the PRISMA guideline for scoping reviews (Tricco et al., 2018). To ensure a comprehensive search of the diversity of literature, the search strategy included three databases as well as reference list searches.

Despite the strengths of this work, there are some notable limitations. Foremost is the lack of data. Across the three data streams we relied on, there is relatively minimal information (i.e., contaminant sources and types, human exposure pathways, health outcomes) on federal contaminated sites to which Indigenous peoples are potentially exposed. Within federal databases, there are 3138 contaminated sites listed without data on contaminants, and a small fraction of these inventoried sites have been the focus of peer-reviewed scientific studies. Despite evidence from federal databases on the presence of hundreds of contaminants (i.e., 440 unique contaminants identified) present at contaminated sites, there is almost no peer-reviewed research on community-specific exposures or health outcomes in the literature. Such a lack of knowledge (i.e., unknown potential health and environmental effects) has been shown to create stress and fear for many Indigenous communities, thereby changing their relationship with the land (Cassady, 2007; Hoover, 2013; Smith et al., 2010; Teufel-Shone et al., 2021).

Furthermore, due to the relatively small and disparate body of knowledge on the issue of contaminated sites and Indigenous peoples in North America, it is difficult to draw strong conclusions on the findings (though our findings are generally consistent). While there has been a legacy of challenges related to contaminated sites for Indigenous peoples, the documentation of federal contaminated sites is relatively recent, with the US Superfund program established in 1980 (Beins, 2015) and the Canadian Contaminated Sites Working Group established in 1995 (Contaminated Sites Management Working Group, 2000). According to our review, peer-reviewed literature available on the topic is dated as early as 1996. In addition, Indigenous peoples across Canada and the US are not a homogenous group, and the findings of this review are not generalizable. However, the nature of a scoping review is that it provides a general overview of current knowledge on the topic and can thus provide some motivation and direction for future research.

This study did not identify all relevant grey literature, but rather sought grey literature iteratively as the review developed per the SYMBALS protocol described in the methods (van Haastrecht et al., 2021). This strategy helped with the management of the breadth of concepts within the broad topic of contaminated sites and Indigenous communities and enabled the gathering of multiple perspectives on a single concept, as opposed to single perspectives on multiple topics. The included literature was reviewed by one reviewer, and this is a major limitation. However, the relatively small body of literature available allowed for the selected articles to be reviewed with a high degree of detail and rigor by one reviewer. Finally, and as elaborated in the methods section (Positionality Statement), we note that this review was conducted by non-Indigenous academics, who have a limited understanding of the experiences of Indigenous peoples and communities. Every attempt was made to acknowledge this positionality and ensure transparency in the process of writing this article, however, this is a major limitation.

### 4.2 Conclusions

This scoping review considered academic research, grey literature, and federal data to better understand the issue of contaminated sites and Indigenous peoples in Canada and the US. In doing so, we found a vast and diverse but also disparate body of information evidencing the contamination of the lands of 815 distinct Indigenous tribes and nations and the presence of 440 different contaminants or contaminant groups. Our first objective was to synthesize information pertaining to the relationship between contaminated sites and Indigenous people, and their land and food systems. Results demonstrated that there is minimal data on the contaminant sources and types, human exposure pathways, health outcomes assessment and management of risks relative to the number of sites on or adjacent to Indigenous lands. Our second objective was to better understand the strategies, challenges, and successes for contaminated sites assessment and management on Indigenous land, and our third objective was to explore the evidence on Indigenous leadership and inclusion in contaminated sites management and research activities.

Overall, our review demonstrated a need for more holistic, upstream, and efficient approaches in the assessment, management, and research of contaminated sites on Indigenous lands. This should include a focus on community-specific approaches to site management and a re-conceptualization of risks related to contaminated sites that privileges Indigenous epistemologies; greater collaboration between networks such as the scientific community, Indigenous communities, and federal governments; and a re-evaluation of current frameworks in which contaminated sites are addressed with Indigenous leadership at the forefront.

## Supporting information

Supplemental Materials Word Document

## Data Availability

Data that was analyzed as part of this scoping review is available as a supplemental Excel file and includes data extraction spreadsheets from the peer-reviewed literature and grey literature included, and original data retrieved from the Canadian Federal Contaminated Sites Inventory and Superfund Sites Data repository, which are publically available online (links provided).

https://www.tbs-sct.gc.ca/fcsi-rscf/numbers-numeros-eng.aspx?qid=872789

https://cumulis.epa.gov/supercpad/cursites/srchsites.cfm

## ACKNOWLEDGEMENTS

We acknowledge funding support to KC from McGill University Faculty of Agricultural and Environmental Sciences (Sustainable Agriculture Fellowship, Graduate Excellence Award) and the PURE CREATE program funded by Natural Sciences and Engineering Research Council of Canada (NSERC). NB’s contribution was supported by the Canada Research Chairs (CRC) Program.

## Notes

**DECLARATION OF CONFLICTS OF INTEREST** The authors have no conflicts of interest to declare

### Competing Interest Statement

The authors have declared no competing interest.

## References

Arquette, M., Cole, M., Cook, K., LaFrance, B., Peters, M., Ransom, J., Sargent, E., Smoke, V., & Stairs, A. (2002, APR). Holistic risk-based environmental decision making: A native perspective. Environmental health perspectives, 110, 259–264. https://doi.org/10.1289/ehp.02110s2259

Arsenault, R., Bourassa, C., Diver, S., McGregor, D., & Witham, A. (2019). Including indigenous knowledge systems in environmental assessments: restructuring the process. Global Environmental Politics, 19(3), 120–132.

Assembly of First Nations. (2001, May 11-13, 2001). After the Mine: Healing Our Lands and Nations-a workshop on abandoned mines The Assembly of First Nations and MiningWatch Canada, Sudbury, Ontario. https://miningwatch.ca/sites/default/files/afn_mwc_workshop_report.pdf

Beins, K. a. L., Stephen. (2015). Superfund: Polluters pay so children can play. https://chej.org/wp-content/uploads/Superfund-35th-Anniversary-Report1.pdf

Bienkowski, B. (2012). Contaminated Culture: Native People Struggle with Tainted Resources. Scientific American.

Blake, J. M., Avasarala, S., Artyushkova, K., Ali, A.-M. S., Brearley, A. J., Shuey, C., Robinson, W. P., Nez, C., Bill, S., & Lewis, J. (2015). Elevated concentrations of U and co-occurring metals in abandoned mine wastes in a northeastern Arizona Native American community. Environmental Science & Technology, 49(14), 8506–8514.

Bland, A. D., Kegler, M. C., Escoffery, C., & Malcoe, L. H. (2005, Jul). Understanding childhood lead poisoning preventive behaviors: the roles of self-efficacy, subjective norms, and perceived benefits. Preventive Medicine, 41(1), 70–78. https://doi.org/10.1016/j.ypmed.2004.10.010

Briffa, J., Sinagra, E., & Blundell, R. (2020, 2020/09/01/). Heavy metal pollution in the environment and their toxicological effects on humans. Heliyon, 6(9), e04691. https://doi.org/https://doi.org/10.1016/j.heliyon.2020.e04691

Brown, T. M., Fisk, A. T., Helbing, C. C., & Reimer, K. J. (2014). Polychlorinated biphenyl profiles in ringed seals (Pusa Hispida) reveal historical contamination by a military radar station in Labrador, Canada. ENVIRONMENTAL TOXICOLOGY AND CHEMISTRY, 33(3), 592–601.

Brown, T. M., Kuzyk, Z. Z. A., Stow, J. P., Burgess, N. M., Solomon, S. M., Sheldon, T. A., & Reimer, K. J. (2013). Effects-based marine ecological risk assessment at a polychlorinated biphenyl-contaminated site in Saglek, Labrador, Canada. ENVIRONMENTAL TOXICOLOGY AND CHEMISTRY, 32(2), 453–467. https://doi.org/https://doi.org/10.1002/etc.2070

Brugge, D., delemos, J. L., & Bui, C. (2007). The Sequoyah Corporation fuels release and the Church Rock spill: unpublicized nuclear releases in American Indian communities. American journal of public health, 97(9), 1595–1600.

Buell, M.-C., Ritchie, D., Ryan, K., & Metcalfe, C. D. (2020). Using Indigenous and Western knowledge systems for environmental risk assessment. Ecological Applications, 30(7), e02146. https://doi.org/https://doi.org/10.1002/eap.2146

Burger, J. (2004a). Recreational rates and future land-use preferences for four Department of Energy sites: consistency despite demographic and geographical differences. ENVIRONMENTAL RESEARCH, 95(2), 215–223.

Burger, J. (2004b). Study of the future land use of a contaminated site: Preferences versus potential use. Remediation Journal: The Journal of Environmental Cleanup Costs, Technologies & Techniques, 14(4), 97–110.

Canadian Council of Ministers of the Environment. (2008). National Classification System for Contaminated Sites-Guidance Document. https://www.ccme.ca/en/res/ncscs_guidance_e.pdf

Canadian Geographic. Metis Settlements and Farms. In Indigenous People’s Atlas of Canada. Canadian Geographic. https://indigenouspeoplesatlasofcanada.ca/article/metis-settlements-and-farms/

Canuel, R., Grosbois, S. B. d., Atikessé, L., Lucotte, M., Arp, P., Ritchie, C., Mergler, D., Chan, H. M., Amyot, M., & Anderson, R. (2006). New Evidence on Variations of Human Body Burden of Methylmercury from Fish Consumption. Environmental health perspectives, 114(2), 302–306. https://doi.org/doi:10.1289/ehp.7857

Cassady, J. (2007). A tundra of sickness: the uneasy relationship between toxic waste, TEK, and cultural survival. Arctic Anthropology, 44(1), 87–97.

Castleden, H., Bennett, E., Lewis, D., & Martin, D. (2017). “ Put It Near the Indians”: Indigenous Perspectives on Pulp Mill Contaminants in Their Traditional Territories (Pictou Landing First Nation, Canada). Progress in community health partnerships: research, education, and action, 11(1), 25–33.

Chan, H. M., Fediuk, K., Batal, M., Sadik, T., Tikhonov, C., Ing, A., & Barwin, L. (2021, 2021/06/01). The First Nations Food, Nutrition and Environment Study (2008–2018)—rationale, design, methods and lessons learned. Canadian Journal of Public Health, 112(1), 8–19. https://doi.org/10.17269/s41997-021-00480-0

Chan, H. M., Singh, K., Batal, M., Marushka, L., Tikhonov, C., Sadik, T., Schwartz, H., & Fediuk, K. (2021). Levels of metals and persistent organic pollutants in traditional foods consumed by First Nations living on-reserve in Canada. Canadian Journal of Public Health, 112(1), 81–96.

Clark, T. (2020). A Comparison of Tribal Sovereignty, Self-Determination, Environmental Justice at the EPA’s Onondaga Lake and Superfund Sites City University of New York]. New York. https://academicworks.cuny.edu/gc_etds/3739/

Contaminated Sites Management Working Group. (2000). Federal approach to contaminated sites. https://www.canada.ca/en/environment-climate-change/services/federal-contaminated-sites/federal-approach.html

Coté, C. (2016). “Indigenizing” food sovereignty. Revitalizing Indigenous food practices and ecological knowledges in Canada and the United States. Humanities, 5(3), 57.

Crown-Indigenous and Northern Affairs Canada. (2021). Evaluation of the Northern Contaminated Sites Program. https://www.rcaanc-cirnac.gc.ca/DAM/DAM-CIRNAC-RCAANC/DAM-AEV/STAGING/texte-text/ev_ncsp21_1628679495032_eng.pdf

Crown-Indigenous and Northern Affairs Canada. (2022). Indigenous Peoples and Communities. Crown-Indigenous and Northern Affairs Canada. Retrieved June 27 from https://www.rcaanc-cirnac.gc.ca/eng/1100100013785/1529102490303

Damman, S., Eide, W. B., & Kuhnlein, H. V. (2008, 2008/04/01/). Indigenous peoples’ nutrition transition in a right to food perspective. Food Policy, 33(2), 135–155. https://doi.org/https://doi.org/10.1016/j.foodpol.2007.08.002

deLemos, J. L., Brugge, D., Cajero, M., Downs, M., Durant, J. L., George, C. M., Henio-Adeky, S., Nez, T., Manning, T., Rock, T., Seschillie, B., Shuey, C., & Lewis, J. (2009, 2009/07/09). Development of risk maps to minimize uranium exposures in the Navajo Churchrock mining district. Environmental Health, 8(1), 29. https://doi.org/10.1186/1476-069X-8-29

Denham, M., Schell, L. M., Deane, G., Gallo, M. V., Ravenscroft, J., & DeCaprio, A. P. (2005, Feb). Relationship of lead, mercury, mirex, dichlorodiphenyldichloroethylene, hexachlorobenzene, and polychlorinated biphenyls to timing of menarche among Akwesasne Mohawk girls. Pediatrics, 115(2), e127–134. https://doi.org/10.1542/peds.2004-1161

Doyle, J. R., Blais, J. M., Holmes, R. D., & White, P. A. (2012, May 1). A soil ingestion pilot study of a population following a traditional lifestyle typical of rural or wilderness areas. SCIENCE OF THE TOTAL ENVIRONMENT, 424, 110–120. https://doi.org/10.1016/j.scitotenv.2012.02.043

Eckert, L. E., Claxton, N. X., Owens, C., Johnston, A., Ban, N. C., Moola, F., & Darimont, C. T. (2020). Indigenous knowledge and federal environmental assessments in Canada: applying past lessons to the 2019 impact assessment act. Facets, 5(1), 67–90.

Ellison, M. (2012). Development of Aboriginal Lands: Successes, risks and environemental concerns respecting contaminated sites Vancouver, BC

Emel, J., & Krueger, R. (2003). Spoken but not Heard: The promise of the precautionary principle for natural resource development. Local Environment, 8, 9–25.

Engwa, G. A., Ferdinand, P. U., Nwalo, F. N., & Unachukwu, M. N. (2019). Mechanism and health effects of heavy metal toxicity in humans. Poisoning in the modern world-new tricks for an old dog, 10.

Environment and Climate Change Canada. (2019, 2019-07-24). Federal Contaminated Sites: Success Stories. Environment and Climate Change Canada. Retrieved February 15 from https://www.canada.ca/en/environment-climate-change/services/federal-contaminated-sites/success-stories.html#kitasoo

EPA. (1996). Third National Tribal Conference on Environmental Management.

EPA. (2011a). EPA Policy on Consultation and Coordination with Indian Tribes. https://www.epa.gov/sites/default/files/2013-08/documents/cons-and-coord-with-indian-tribes-policy.pdf

EPA. (2011b). This is Superfund A community Guide to EPA’s Superfund Program. http://www7.nau.edu/itep/main/hazsubmap/docs/CERCLA/ThisIsSuperfundACommunityGuideT oEPAsSuperfundProgram9.13.11.pdf

EPA. (2015). A Decade of Tribal Environmental Health Research: Results and Impacts from EPA’s Extramural Grants and Fellowship Programs. https://www.epa.gov/sites/default/files/2015-08/documents/results-impacts.pdf

EPA. (2017). Revised HRS Final Rule, 40 CFR 300. https://semspub.epa.gov/work/HQ/100002489.pdf

EPA. (2020). Population Surrounding 1857 Superfund Remedial Sites. https://outlook.office.com/mail/inbox/id/AAQkADAyNWViYjc3LTI3OGQtNDRjOC04OWU3LTExMjU2Zjg3NzBlZgAQAErw9YWvUtBNqV%2F%2FtjXe5gY%3D?realm=mcgill.ca&path=/mail/search

EPA. (2021a). CIMC Listing United States Environmental Protection Agency. https://cimc.epa.gov/ords/cimc/f?p=CIMC:LIST

EPA. (2021b). Cleanups in My Community (CIMC)- Cleanups and Grants Listings Page https://cimc.epa.gov/ords/cimc/f?p=CIMC:LIST

EPA. (2021c, September 28, 2021). Contaminated Land. United States Environmental Protection Agency. https://www.epa.gov/report-environment/contaminated-land#roe-indicators

EPA. (2021d). Search Superfund Site Information https://cumulis.epa.gov/supercpad/cursites/srchsites.cfm

EPA. (2022a). EPA ExpoBox [Database]. EPA Expobox. https://cfpub.epa.gov/ncea/risk/expobox/efhTableSearch.cfm

EPA. (2022b). Land Cleanup Funding Authorities Available to Tribal Governments. Retrieved June 14 from https://www.epa.gov/tribal-lands/land-cleanup-funding-authorities-available-tribal-governments-0

EPA. (2022c). Land Cleanup Funding Authorities Available to Tribal Governments. Retrieved May 27 2022 from https://www.epa.gov/tribal-lands/land-cleanup-funding-authorities-available-tribal-governments-0

EPA. (2022d). Risk Management. Retrieved May 30 from https://www.epa.gov/risk/risk-management

Ferguson, K. (2021). Indigenous Environmental health research secures $1.3M grant. Western News. https://news.westernu.ca/2021/08/indigenous-environmental-health-research-secures-1-3m-grant/

Fernández-Llamazares, Á., Garteizgogeascoa, M., Basu, N., Brondizio, E. S., Cabeza, M., Martínez-Alier, J., McElwee, P., & Reyes-García, V. (2020). A State-of-the-Art Review of Indigenous Peoples and Environmental Pollution. Integrated Environmental Assessment and Management, 16(3), 324–341. https://doi.org/https://doi.org/10.1002/ieam.4239

Fitzgerald, E. F., Brix, K. A., Deres, D. A., Hwang, S.-A., Bush, B., Lambert, G., & Tarbell, A. (1996). Polychlorinated biphenyl (PCB) and dichlorodiphenyl dichloroethylene (DDE) exposure among Native American men from contaminated Great Lakes fish and wildlife. Toxicology and industrial health, 12(3-4), 361–368.

Fitzgerald, E. F., Hwang, S.-A., Langguth, K., Cayo, M., Yang, B.-Z., Bush, B., Worswick, P., & Lauzon, T. (2004, 2004/02/01/). Fish consumption and other environmental exposures and their associations with serum PCB concentrations among Mohawk women at Akwesasne. ENVIRONMENTAL RESEARCH, 94(2), 160–170. https://doi.org/https://doi.org/10.1016/S0013-9351(03)00133-6

Flett, L., McLeod, C. L., McCarty, J. L., Shaulis, B. J., Fain, J. J., & Krekeler, M. P. S. (2021, 2021/03/01/). Monitoring uranium mine pollution on Native American lands: Insights from tree bark particulate matter on the Spokane Reservation, Washington, USA. ENVIRONMENTAL RESEARCH, 194, 110619. https://doi.org/https://doi.org/10.1016/j.envres.2020.110619

Gailus, J. (2013). Management of Contaminated Sites on Indian Reserve Lands Site Remediation in British Colombia Conference, Vancouver. https://www.dgwlaw.ca/wp-content/uploads/2014/12/Site_Remediation_Conference_Paper.pdf

Gallo, M. (2011). From wood treatment to unequal treatment: The story of the St. Regis Superfund site. Law & Ineq., 29, 175.

Garvin, E. M., Bridge, Cas F., Garvin, Meredith S. (2018). Edible wild plants growing in contaminated floodplains: implications for the issuance of tribal consumption advisories within the Grand Lake watershed of northeastern Oklahoma, USA. Environmental Geochemistry and Health, 40(3), 999–1025. https://doi.org/http://dx.doi.org/10.1007/s10653-017-9960-3

Goncharov, A., Haase, R. F., Santiago-Rivera, A., Morse, G., McCaffrey, R. J., Rej, R., & Carpenter, D. O. (2008, 2008/02/01/). High serum PCBs are associated with elevation of serum lipids and cardiovascular disease in a Native American population. ENVIRONMENTAL RESEARCH, 106(2), 226–239. https://doi.org/https://doi.org/10.1016/j.envres.2007.10.006

Gover, L. (2007). Twenty Years Later-Tribes and the Superfund Program. Natural Resources & Environment, 21(3), 48–53.

Indian Act, (1985). https://laws-lois.justice.gc.ca/eng/acts/i-5/fulltext.html

Government of Canada. (2012). Report of the Commissioner of the Environmental and Sustainable Development-Federal Contaminated Sites and Their Impacts. http://www.oagbvg.gc.ca/internet/English/parl_cesd_201205_03_e_36775.html#hd5b,

Government of Canada. (2017). Committee report-November 16–17, 2016. . https://www.canada.ca/en/health-canada/services/chemical-substances/chemicals-management-plan/science-committee/meeting-records-reports/committee-report-november-16-17-2016.html

Government of Canada. (2019). Action plan for contaminated sites. Government of Canada. Retrieved May 24 from https://www.canada.ca/en/environment-climate-change/services/federal-contaminated-sites/action-plan.html

Government of Canada. (2021). Federal Contaminated Sites Inventory. https://www.tbs-sct.gc.ca/fcsi-rscf/home-accueil-eng.aspx

Government of Northwest Territories. (2021). Human Health Risk Assessment for Legacy Arsenic Contamination Around Yellowknife-Plain Language Summary. https://www.enr.gov.nt.ca/sites/enr/files/resources/arsenic_human_health_risk_assessment_summary_june_2021_0.pdf

Gunn K, O. N. C. (2021). Indigenous Law and Canadian Courts. First Peoples’ Law. https://www.firstpeopleslaw.com/public-education/blog/indigenous-law-canadian-

Haalboom, B., & Natcher, D. C. (2012). The power and peril of“ vulnerability”: Approaching community labels with caution in climate change research. ARCTIC, 319–327.

Haddaway, N. R., Collins, A. M., Coughlin, D., & Kirk, S. (2015). The role of Google Scholar in evidence reviews and its applicability to grey literature searching. PLoS ONE, 10(9), e0138237.

Hansen, T. (2018). Kill the Land, Kill the People: There Are 532 Superfund Sites in Indian Country! Indian Country Today.

Harper, B. (2007). Traditional Tribal Subsistence Exposure Scenario and Risk Assessment Guidance Manual. https://health.oregonstate.edu/sites/health.oregonstate.edu/files/research/pdf/tribal-grant/exposure_scenario_and_risk_guidance_manual_v2.pdf

Harper, B., Harding, A., Harris, S., & Berger, P. (2012). Subsistence exposure scenarios for tribal applications. Human and Ecological Risk Assessment: An International Journal, 18(4), 810–831.

Harper, B. L., Flett, B., Harris, S., Abeyta, C., & Kirschner, F. (2002). The Spokane Tribe’s multipathway subsistence exposure scenario and screening level RME. RISK ANALYSIS, 22(3), 513–526.

Harris, S. G., & Harper, B. L. (1997, Dec). A Native American exposure scenario. RISK ANALYSIS, 17(6), 789–795. https://doi.org/10.1111/j.1539-6924.1997.tb01284.x

Health Canada. (2010). A Guide to Involving Aboriginal Peoples in Contaminated Sites Managament. https://publications.gc.ca/collections/collection_2011/sc-hc/H128-1-10-628-eng.pdf

Health Canada. (2018). Guidance for evaluating human health impacts in environmental assessment: country foods.

Holifield, R. (2012). Environmental Justice as Recognition and Participation in Risk Assessment: Negotiating and Translating Health Risk at a Superfund Site in Indian Country. ANNALS OF THE ASSOCIATION OF AMERICAN GEOGRAPHERS, 102(3), 591–613.https://doi.org/10.1080/00045608.2011.641892

Hoover, E. (2013). Cultural and health implications of fish advisories in a Native American community. Ecological Processes, 2(1), 1–12.

Hoover, E. (2016). “ We’re not going to be guinea pigs;” Citizen Science and Environmental Health in a Native American Community. Journal of Science Communication, 15(1), A05.

Hund, L., Bedrick, E. J., Miller, C., Huerta, G., Nez, T., Ramone, S., Shuey, C., Cajero, M., & Lewis, J. (2015). A Bayesian framework for estimating disease risk due to exposure to uranium mine and mill waste on the Navajo Nation. Journal of the Royal Statistical Society: Series A (Statistics in Society), 178(4), 1069–1091.

Hwang, S.-A., Yang, B.-Z., Fitzgerald, E. F., Bush, B., & Cook, K. (2001, 2001/07/01). Fingerprinting PCB patterns among Mohawk women. Journal of Exposure Science & Environmental Epidemiology, 11(3), 184–192. https://doi.org/10.1038/sj.jea.7500159

Hykin, J. B. (2016, September 21, 2016). Contaminated Sites on First Nations Lands 2016 Site Remediation Conference, Vancouver, BC. http://www.woodwardandcompany.com/wp-content/uploads/pdfs/2016-09-20-Contaminated_Sites_on_First_Nation_Lands-Final.pdf

Indian Country Today. (2013). Mohawks Say EPA Alcoa-Superfund Cleanup Plan Falls Short. Indian Country Today. https://indiancountrytoday.com/archive/mohawks-say-epa-alcoa-superfund-cleanup-plan-falls-short

Indigenous and Northern Affairs Canada. (2016). Evaluation of the Contaminated Sites On-Reserve (South of the 60th Parallel) Program. https://www.rcaanc-cirnac.gc.ca/DAM/DAM-CIRNAC-RCAANC/DAM-AEV/STAGING/texte-text/ev_css60_1511896979296_eng.pdf

Indigenous Services Canada. (2020). Annual Report to Parliament https://www.sac-isc.gc.ca/DAM/DAM-ISC-SAC/DAM-TRNSPRCY/STAGING/texte-text/annual-report-parliament-arp-report2020_1648059621383_eng.pdf

Indigenous Services Canada. (2021). First Nations Environmental Contaminants Program. Retrieved January 31 from https://www.sac-isc.gc.ca/eng/1583779185601/1583779243216

Jonasson, M. E., Spiegel, S. J., Thomas, S., Yassi, A., Wittman, H., Takaro, T., Afshari, R., Markwick, M., & Spiegel, J. M. (2019, 2019/12/01). Oil pipelines and food sovereignty: threat to health equity for Indigenous communities. Journal of Public Health Policy, 40(4), 504–517. https://doi.org/10.1057/s41271-019-00186-1

Jones, N. M., Rachel; Ramirez, Roberto; Rios-Vargas, Merarys. (2021, August 12, 2021). 2020 Census Illuminates Racial and Ethic Composition of the Country. United States Census Bureau. https://www.census.gov/library/stories/2021/08/improved-race-ethnicity-measures-reveal-united-states-population-much-more-multiracial.html

Joseph, B. (2020, July 4). What is the seventh generation principle? https://www.ictinc.ca/blog/seventh-generation-principle#:~:text=The%20Seventh%20Generation%20Principle%20is,seven%20generations%20into%20the%20future.

Kegler, M. C., & Malcoe, L. H. (2004). Results from a lay health advisor intervention to prevent lead poisoning among rural Native American children. American journal of public health, 94(10), 1730–1735.

Kegler, M. C., Malcoe, L. H., & Fedirko, V. (2010). Primary prevention of lead poisoning in rural native American children: behavioral outcomes from a community-based intervention in a former mining region. Family and Community Health, 32–43.

Kegler, M. C., Rigler, J., & Ravani, M. K. (2010). Using network analysis to assess the evolution of organizational collaboration in response to a major environmental health threat. Health Education Research, 25(3), 413–424. https://doi.org/10.1093/her/cyq022

Kent, T. (2016). Tribal-Led Cleanup Activities at the Tar Creek Superfund Site Tribal Lands Environmental Forum, https://www7.nau.edu/itep/main/iteps/ORCA/6354_ORCA.pdf

Kerfoot, W. C., Urban, N., Jeong, J., MacLennan, C., & Ford, S. (2020, 2020/10/01/). Copper-rich “Halo” off Lake Superior’s Keweenaw Peninsula and how Mass Mill tailings dispersed onto tribal lands. JOURNAL OF GREAT LAKES RESEARCH, 46(5), 1423–1443. https://doi.org/https://doi.org/10.1016/j.jglr.2020.07.004

Koch, I., Dee, J., House, K., Sui, J., Zhang, J., McKnight-Whitford, A., & Reimer, K. J. (2013, 2013/04/01/). Bioaccessibility and speciation of arsenic in country foods from contaminated sites in Canada. SCIENCE OF THE TOTAL ENVIRONMENT, 449, 1–8. https://doi.org/https://doi.org/10.1016/j.scitotenv.2013.01.047

Krewski, D., Acosta, D., Jr., Andersen, M., Anderson, H., Bailar, J. C., 3rd, Boekelheide, K., Brent, R., Charnley, G., Cheung, V. G., Green, S., Jr., Kelsey, K. T., Kerkvliet, N. I., Li, A. A., McCray, L., Meyer, O., Patterson, R. D., Pennie, W., Scala, R. A., Solomon, G. M., Stephens, M., Yager, J., & Zeise, L. (2010). Toxicity testing in the 21st century: a vision and a strategy. Journal of toxicology and environmental health. Part B, Critical reviews, 13(2-4), 51–138. https://doi.org/10.1080/10937404.2010.483176

Landrigan, P. J., Fuller, R., Acosta, N. J. R., Adeyi, O., Arnold, R., Basu, N., Baldé, A. B., Bertollini, R., Bose-O’Reilly, S., Boufford, J. I., Breysse, P. N., Chiles, T., Mahidol, C., Coll-Seck, A. M., Cropper, M. L., Fobil, J., Fuster, V., Greenstone, M., Haines, A., Hanrahan, D., Hunter, D., Khare, M., Krupnick, A., Lanphear, B., Lohani, B., Martin, K., Mathiasen, K. V., McTeer, M. A., Murray, C. J. L., Ndahimananjara, J. D., Perera, F., Potočnik, J., Preker, A. S., Ramesh, J., Rockström, J., Salinas, C., Samson, L. D., Sandilya, K., Sly, P. D., Smith, K. R., Steiner, A., Stewart, R. B., Suk, W. A., van Schayck, O. C. P., Yadama, G. N., Yumkella, K., & Zhong, M. (2018). The Lancet Commission on pollution and health. The Lancet, 391(10119), 462–512. https://doi.org/10.1016/S0140-6736(17)32345-0

LeClerc, E., & Keeling, A. (2015, 2015/01/01/). From cutlines to traplines: Post-industrial land use at the Pine Point mine. The Extractive Industries and Society, 2(1), 7–18. https://doi.org/https://doi.org/10.1016/j.exis.2014.09.001

Lewis, J., Gonzales, M., Burnette, C., Benally, M., Seanez, P., Shuey, C., Nez, H., Nez, C., & Nez, S. (2015). Environmental exposures to metals in native communities and implications for child development: basis for the Navajo Birth Cohort Study. Journal of Social Work in Disability & Rehabilitation, 14(3-4), 245–269.

Lewis, J., Hoover, J., & MacKenzie, D. (2017, 2017/06/01). Mining and Environmental Health Disparities in Native American Communities. Current environmental health reports, 4(2), 130–141. https://doi.org/10.1007/s40572-017-0140-5

Mahood, Q., Van Eerd, D., & Irvin, E. (2014). Searching for grey literature for systematic reviews: challenges and benefits. Research synthesis methods, 5(3), 221–234.

McAuley, C., & Knopper, L. D. (2011, 2011/06/08). Impacts of traditional food consumption advisories: Compliance, changes in diet and loss of confidence in traditional foods. Environmental Health, 10(1), 55. https://doi.org/10.1186/1476-069X-10-55

Meltzer, G., Avenbuan, O., Wu, F., Shah, K., Chen, Y., Mann, V., & Zelikoff, J. T. (2020, 2020/12/01). The Ramapough Lunaape Nation: Facing Health Impacts Associated with Proximity to a Superfund Site. Journal of Community Health, 45(6), 1196–1204. https://doi.org/10.1007/s10900-020-00848-2

Michelsen, T. (2010). Superfund on Tribal Lands: Issues, Challenges, and Solutions. https://clu-in.org/conf/tio/NARPMPresents1_101211/Superfund-on-Tribal-Lands.pdf

Middleton, B. R., Talaugon, S., Young, T. M., Wong, L., Fluharty, S., Reed, K., Cosby, C., & Myers, R. (2019). Bi-directional learning: identifying contaminants on the Yurok Indian reservation. INTERNATIONAL JOURNAL OF ENVIRONMENTAL RESEARCH AND PUBLIC HEALTH, 16(19), 3513.

Moore-Nall, A. (2015). The legacy of uranium development on or near Indian reservations and health implications rekindling public awareness. Geosciences, 5(1), 15–29.

Munn, Z., Peters, M. D., Stern, C., Tufanaru, C., McArthur, A., & Aromataris, E. (2018). Systematic review or scoping review? Guidance for authors when choosing between a systematic or scoping review approach. BMC medical research methodology, 18(1), 1–7.

National Conference of State Legislators. (2020). Federal and State Recognized Tribes. National Conference of State Legislators. Retrieved June 27 from https://www.ncsl.org/legislators-staff/legislators/quad-caucus/list-of-federal-and-state-recognized-tribes.aspx#State

National Institute of Environmental Health Sciences. (2022). Superfund Research Program. https://www.niehs.nih.gov/research/supported/centers/srp/index.cfm

Native Land Interactive Map. (2021). Retrieved November 12 from https://native-land.ca/

Nolan, C. G. (2009). Risk assessment and orphaned/abandoned mines in Canada—What role do aboriginal communities play in risk assessment? Integrated Environmental Assessment and Management, 5(3), 486–487.

OECD. (2020). Linking Indigenous Communities with Regional Development in Canada https://doi.org/doi:doi:https://doi.org/10.1787/fa0f60c6-en

Parsons, M., Fisher, K., & Crease, R. P. (2021). Environmental Justice and Indigenous Environmental Justice. Decolonising Blue Spaces in the Anthropocene: Freshwater management in Aotearoa New Zealand, 39-73. https://doi.org/10.1007/978-3-030-61071-5_2

Peters, M. D., Godfrey, C. M., Khalil, H., McInerney, P., Parker, D., & Soares, C. B. (2015). Guidance for conducting systematic scoping reviews. JBI Evidence Implementation, 13(3), 141–146.

Reading, C., & Wien, F. (2009). Health Inequities and Social Determinants of Aboriginal Peoples’ Health. https://www.ccnsa-nccah.ca/docs/determinants/RPT-HealthInequalities-Reading-Wien-EN.pdf

Reid, A. J., Eckert, L. E., Lane, J. F., Young, N., Hinch, S. G., Darimont, C. T., Cooke, S. J., Ban, N. C., & Marshall, A. (2021). “Two-Eyed Seeing”: An Indigenous framework to transform fisheries research and management. Fish and Fisheries, 22(2), 243–261.

Rock, T., Camplain, R., Teufel-Shone, N. I., & Ingram, J. C. (2019). Traditional sheep consumption by Navajo people in Cameron, Arizona [Article]. INTERNATIONAL JOURNAL OF ENVIRONMENTAL RESEARCH AND PUBLIC HEALTH, 16(21), Article 4195. https://doi.org/10.3390/ijerph16214195

Samuel-Nakamura, C., Robbins, W. A., & Hodge, F. S. (2017). Uranium and associated heavy metals in Ovis aries in a mining impacted area in Northwestern New Mexico. INTERNATIONAL JOURNAL OF ENVIRONMENTAL RESEARCH AND PUBLIC HEALTH, 14(8), 848.

Sandlos, J., & Keeling, A. (2016, APR). Aboriginal communities, traditional knowledge, and the environmental legacies of extractive development in Canada. EXTRACTIVE INDUSTRIES AND SOCIETY-AN INTERNATIONAL JOURNAL, 3(2), 278–287. https://doi.org/10.1016/j.exis.2015.06.005

Sandlos, J., & Keeling, A. (2016). Toxic legacies, slow violence, and environmental injustice at Giant Mine, Northwest Territories. Northern Review(42), 7–21-27–21.

Sarkar, A., Wilton, D. H. C., Fitzgerald, E., Sharma, A., Sharma, A., & Sathya, A. J. (2019, Apr). Environmental impact assessment of uranium exploration and development on indigenous land in Labrador (Canada): a community-driven initiative. Environ Geochem Health, 41(2), 939–949. https://doi.org/10.1007/s10653-018-0191-z

Schmitt, C. J., Brumbaugh, W. G., Linder, G. L., & Hinck, J. E. (2006, 2006/10/01). A Screening-Level Assessment of Lead, Cadmium, and Zinc in Fish and Crayfish from Northeastern Oklahoma, USA. Environmental Geochemistry and Health, 28(5), 445–471. https://doi.org/10.1007/s10653-006-9050-4

Sistili, B., Metatawabin, M., Iannucci, G., & Tsuji, L. J. (2006). An Aboriginal perspective on the remediation of mid-Canada radar line sites in the Subarctic: A partnership evaluation. ARCTIC, 142–154.

Smith, K., Luginaah, I., & Lockridge, A. (2010). Contaminated’therapeutic landscape: the case of the Aamjiwnaang First Nation in Ontario, Canada. Geography Research Forum,

Teufel-Shone, N. I., Chief, C., Richards, J. R., Clausen, R. J., Yazzie, A., Begay, M. A., Jr., Lothrop, N., Yazzie, J., Begay, A. B., Beamer, P. I., & Chief, K. (2021, Sep 6). Development of a Culturally Anchored Qualitative Approach to Conduct and Analyze Focus Group Narratives Collected in Diné (Navajo) Communities to Understand the Impacts of the Gold King Mine Spill of 2015. Int J Environ Res Public Health, 18(17). https://doi.org/10.3390/ijerph18179402

Tribal Superfund Working Group. (2022). Tribal Superfund Working Group. Retrieved January 31 from https://triballands.org/tlac/building-relationships/tribal-superfund-working-group/

Tricco, A. C., Lillie, E., Zarin, W., O’Brien, K. K., Colquhoun, H., Levac, D., Moher, D., Peters, M. D., Horsley, T., & Weeks, L. (2018). PRISMA extension for scoping reviews (PRISMA-ScR): checklist and explanation. Annals of internal medicine, 169(7), 467–473.

Tsuji, L. J., Wainman, B. C., Weber, J.-P., Sutherland, C., Katapatuk, B., & Nieboer, E. (2005). Protecting the health of First Nation personnel at contaminated sites: A case study of Mid-Canada Radar Line Site 050 in northern Canada. ARCTIC, 233–240.

United States Government Accountability Office. (2020). EPA Grants to Tribes-Additional Actions Needed to Effectively Address Tribal Environmental Concerns. https://www.gao.gov/assets/gao-21-150.pdf

US Government Accountability Office. (2019). Superfund: EPA Should Improve the Reliability of Data on National Priorities List Sites Affecting Indian Tribes. https://www.gao.gov/products/gao-19-123

US Government Publishing Office. (2016). Examining EPA’s unacceptable response to Indian Tribes. https://www.govinfo.gov/content/pkg/CHRG-114shrg20899/html/CHRG-114shrg20899.htm

van Haastrecht, M., Sarhan, I., Yigit Ozkan, B., Brinkhuis, M., & Spruit, M. (2021, 2021-May-28). >SYMBALS: A Systematic Review Methodology Blending Active Learning and Snowballing [Original Research]. Frontiers in Research Metrics and Analytics, 6. https://doi.org/10.3389/frma.2021.685591

Wang, Z., Walker, G. W., Muir, D. C. G., & Nagatani-Yoshida, K. (2020, 2020/03/03). Toward a Global Understanding of Chemical Pollution: A First Comprehensive Analysis of National and Regional Chemical Inventories. Environmental Science & Technology, 54(5), 2575–2584. https://doi.org/10.1021/acs.est.9b06379

Wiseman, C. L., & Gobas, F. A. (2002). Balancing risks in the management of contaminated first nations fisheries. International journal of environmental health research, 12(4), 331–342.

Woolford, J. (2017). Consideration of Tribal Treaty Rights and Traditional Ecological Knowledge in the Supefund Remedial Program. https://semspub.epa.gov/work/HQ/500024668.pdf

