## Supplemental Materials Word Document for "Contaminated Sites and Indigenous Peoples in Canada and the United States: A Scoping Review"

**Declaration of conflicts of interest:** The authors have no conflicts of interest to declare

Table of Contents:

- Supplemental Materials, Glossary of Key Terms
- Table S1: Grey Literature Search Terms
- Table S2: Key government webpages used to supplement and interpret the results
- Table S3: Summary of Grey Literature
- Table S4: Studies Measuring Contaminant Concentrations in Environmental Samples
- Table S5: Studies on contaminants in food
- Table S6: Studies on Human Health Outcomes
- Table S7: Peer-reviewed articles that collected primary qualitative data on the impacts of contaminated sites
- Table S8: Contaminated site processes (Canada and US)
- Table S9: Peer-reviewed literature on Health Risk Assessment
- Table S10: Peer-reviewed literature using secondary data (i.e.: did not collect primary data (i.e.: reviews, discussions, and case studies))
- Table S11: Peer-reviewed articles focused on risk management strategies
- Table S12: Peer-reviewed studies collecting primary data on post-remediation land use
- Table S13: Peer-reviewed articles that collected primary data Indigenous Inclusion and Collaboration
- Table S14: Number of peer-reviewed articles collecting data per contaminant media
- Table S15: Indigenous Communities of Focus in Peer-reviewed Literature
- Table S16: Contaminated site processes (Canada and US)
- Figure S1**:** Governmental Programming and Legislation Chart
- Figure S2: Screen Shots of Search Strategy

### **Supplemental Materials, Glossary of Key Terms**

**Contaminated Site (Government of Canada):** A site at which substances occur at concentrations (1) above background (normally occurring) levels and pose or are likely to pose an immediate or long term hazard to human health or the environment, or (2) exceeding levels specified in policies and regulations ^1^.

**Federal Contaminated Sites:** Federal contaminated sites are located on land owned or leased by the federal government, or on land where the federal government has accepted responsibility for the contamination ^2^.

**Federal Contaminated Sites Inventory (FCSI):** includes information on all known and suspected contaminated sites under the custodianship of federal departments, agencies and consolidated Crown corporations. It also includes non-federal contaminated sites for which the Government of Canada has accepted some or all financial responsibility. It does not include sites where contamination has been caused by, and which are under the control of, enterprise Crown corporations, private individuals, firms or other levels of government ^1^.

**Canadian Environmental Protection Act (CEPA):** An Act with the primary purpose of pollution prevention and the protection of the environment and human health to contribute to sustainable development. This includes activities related to the assessment and management of risks from chemicals, polymers and living organisms; programs related to air and water pollution, hazardous waste, air pollutant and greenhouse gas emissions, ocean disposal and environmental emergencies ^3^.

**Federal Contaminated Sites Action Plan (FCSAP):** A 15-year program established in 2005, with funding of $4.54 billion from the Government of Canada. The program was renewed for another 15 years (2020 to 2034) with $1.16 billion announced in Budget 2019 for the first five years (Phase IV, 2020 to 2024). The objective of FCSAP is to reduce environmental and human health risks from known federal contaminated sites and associated federal financial liabilities, while focusing on the highest priority sites ^4^.

**Human exposure Under Control:** “The Human Exposure Under Control Environmental Indicator is a Government Performance and Results Act (GPRA) measure used by EPA to document whether contamination levels at a site fall within the levels specified by EPA as safe, or if they do not, whether adequate controls are in place to prevent human exposure to contamination”. ^5^

**Bill S-5:** the most recent bill amending CEPA, introduced on February 9, 2022. The bill focuses on recognizing a right to a healthy environment as provided under CEPA and strengthening Canada’s chemicals management regime ^6^.

**Superfund/Comprehensive Environmental Response, Compensation, and Liability Act (CERCLA):** Enacted by congress in 1980, creating a tax on the chemical and petroleum industries and providing broad federal authority to respond directly to releases or threatened releases of hazardous substances that may endanger public health or the environment ^7^.

**Brownfields:** Abandoned, idle or underutilized commercial or industrial properties where past actions have caused environmental contamination, but which still have potential for redevelopment or other economic opportunities. Brownfields are typically located in urban areas. FCSAP may contribute to restore federal brownfields for future use, if sites meet the conditions that make them eligible for program funding ^2^.

**Table S1**: Grey Literature Search Terms

Complete list of search terms used to identify grey literature

| Group | Terms | |
| --- | --- | --- |
| Group 1: Indigenous People | - Indigenous People - Indigenous Communities - Native American - Inuit - Metis - First Nations - Aboriginal - Traditional Food | - Country Food - Tribal Land - Tribe - American Indian - Canadian Indian - Reserve land - Indian Country |
| Group 2: Contaminated Sites | - Contaminated Site - Contaminated Land - Superfund - National Priorities List - Brownfield - Cleanups |  |

**Table S2:** Key government webpages used to supplement and interpret the results

| Government Webpages | | |
| --- | --- | --- |
| **Governing Body** | **Title** | **Key Findings** |
| US EPA | Remedial Program in Indian Country | Outlines basic information on the role of and consultation with tribes in the context of Superfund sites |
| Tribal Lands Assistance Center (Institute for Tribal Professionals) | Superfund Tribal Case Studies | A repository of case studies with the purpose of helping tribal professionals navigate cleanup and remediation efforts and to network with other tribes affected by Superfund sites |
| US EPA | Tribal Lands Cleanup and Spill Prevention Programs | Describes superfund and non-superfund contaminated site programming related to sites on or near tribal lands |
| Indigenous Services Canada | Contaminated Sites Management Program | Describes programming through which contaminated sites on ‘inhabited reserves’ are addressed.  The CSMP is an initiative to identify and document environmental problems on reserve land, with a goal to “reduce crown liabilities” and “improve living conditions on reserves” |
| Indigenous Services Canada | First Nations Environmental Contaminants Program | Describes the FNECP, a funding program through which First Nations can apply to assist them with assessing contamination in their communities |
| Government of Canada | Federal Contaminated Sites: Success Stories | Catalogs case studies of successful contaminated site remediation, including examples of contaminated sites on reserve land including Kitasoo, British Colombia. |
| US EPA | Cleanup and Spill Prevention: Laws and Regulations | Outlines federal laws and land protection laws that may be relevant to tribal governments in relation to contaminated sites |

**Table S3:** Grey literature summary charts

| Government Documents | | | | | | | |
| --- | --- | --- | --- | --- | --- | --- | --- |
| **Author** | **Governing Body** | | | **Title** | | | **Key Findings** |
| US Government Accountability Office | US EPA | | | Superfund: EPA Should Improve the Reliability of Data on National Priorities List Sites Affecting Indian Tribes | | | - EPA does not have reliable, accurate and complete data identifying Superfund Sites located on Tribal lands - EPA does not have a regular review process for this data - EPA’s guidance for determining whether a site has Native American interest is unclear, and there is no reliable data on consultations with tribes |
| Evaluation, Performance, and Review Branch Audit and Evaluation Sector | Indigenous and Northern Affairs Canada | | | Evaluation of the Contaminated  Sites On-Reserve (South of the  60th Parallel) Program | | | - Determined that CSOR has a beneficial impact overall, while challenges include and overall lack of funding to meet the continued demand of new sites, issues enforcing environmental protections on-reserve, and legislative gaps. |
| Michelsen, Teresa  Mediation Solutions | US EPA | | | Superfund on Tribal Lands: Issues, Challenges, and Solutions- Assessment Report | | | - Superfund on tribal lands have additional complexities such as technical, historic, cultural, legal, and jurisdictional challenges - suggested future work including better definitions of consultation, integrating meaningful consultation into practice, improved training for EPA staff, better risk assessment models incorporating tribal land-use scenarios |
| Woolford, James  Office of Superfund Remediation and Technology Innovation | US EPA | | | Consideration of Tribal Treaty Rights and Traditional Ecological Knowledge in the  Superfund Remedial Program | | | - Describes how and when Traditional Ecological Knowledge (TEK) should be considered in Superfund Remedial Program Processes. - Notes that “TEK will not be the sole determining factor” in EPA decision making. |
| U.S. Government Publishing Office | US EPA | | | Examining EPA’s Unacceptable Response to Indian Tribes | | | - Describes proceedings following the Gold King Mine Blowout, which involved toxic wastewater spill into cement creek which affected thousands of Navajo Nation members - Nation members and others expressed concern that EPA had an inadequate response to assessing and cleaning the site |
| Scientific Consulting Group, Inc. | US EPA | | | A Decade of Tribal Environmental Health Research: Results and Impacts from EPA’s Extramural Grants and Fellowship Programs | | | - Research on subsistence foods and water resources- research projects primarily resulting in advisories, including mapping, risk avoidance using GIS - One research project looked at an inexpensive, easy-to-use technology from an Indigenous material to remove contaminants from groundwater |
| News Articles | | | | | | | |
| **Author, year, journal** | | | **Title** | | | **Description** | |
| Indian Country Today | | | Mohawks Say EPA Alcoa-Superfund Cleanup Plan Falls Short | | | Article describes that EPA's remediation decision was not seen as adequate by Mohawk members (i.e.: the contaminated river was not cleaned to a sufficient level)  Additionally, the period available for comments from the community was seen as too short for community members to express all opinions | |
| Hansen, Terri (2018, Indian Country Today) | | | Kill the Land, Kill the People: There Are 532 Superfund Sites in Indian Country! | | | Discusses the disproportionate impact of Superfund sites on Native Americans and describes the potential risks associated with these sites | |
| Brian Bienkowski (2012, Scientific American) | | | Contaminated Culture: Native People Struggle with Tainted Resources | | | News article describing physical, environmental and cultural impacts of contaminated sites on Indigenous people | |
| Conference Materials | | | | | | | |
| **Author (year)** | | | **Title** | | | **Description** | |
| Hykin, J (2016) | | | Contaminated Sites on First Nation Lands | | | - Discusses legislative regimes governing environmental regulation on First Nations lands, and how these impact contaminated sites management | |
| Ellison, M (2012) | | | Development of Aboriginal Lands: Successes, risks and environmental concerns respecting contaminated sites | | | - Describes key challenges and successes in the management of Federal contaminated sites on reserve land in Canada - Attests that Federal government remains the power holder in environmental decision-making and over-rides tribal governments - Overall different goals exist between First Nations and Government of Canada | |
| Gailus, Deven (2013) | | | Management of Contaminated Sites on Indian Reserve Lands | | | - Outlines legal regime for contaminated sites on “Indian Reserve Lands” in Canada, highlighting the barriers that exist and the various options for dealing with contaminated sites from a legal viewpoint | |
| Kent, T (2016) | | | Tribal-Led Cleanup Activities at the Tar Creek Superfund Site | | | - Presentation as part of the Tribal Lands Environmental Forum, Tim Kent of the Quapaw Tribe of Oklahoma describes the site and Tribe-led remedial action activities | |
| Journal Articles (Non-Peer Reviewed) | | | | | | | |
| **Author** | | | **Title** | | | **Description** | |
| Castleden, Bennett, Lewis and Martin (2017) | | | “Put It Near the Indians": Indigenous Perspectives on Pulp Mill Contaminants in Their Traditional Territories (Pictou Landing First Nation, Canada). | | | - Used narrative interviews to conceptualize community well-being in the context of environment and human health connections related to pollution of a tidal estuary - Shows a close connection between Mi’kmaw livelihood, local ecologies, and health and well-being | |
| Lewis J, Gonzales M, Burnette C, et al. (2015) | | | Environmental Exposures to Metals in Native Communities and Implications for Child Development: Basis for the Navajo Birth Cohort Study. Journal of Social Work in Disability and Rehabilitation | | | - describes existing evidence on developmental disability risk in Native American children related to the abandoned mines in the Western United States. Numerous studies have linked low-level metals exposure with birth defects and developmental delays. Concern has emerged among tribal populations that metal exposure from abandoned mines might threaten development of future generations. | |
| Gallo, M (2011) | | | From Wood Treatment to Unequal Treatment: The Story of the St. Regis Superfund Site | | | - Describes environmental injustice and legal dynamics related to the St. Regis Superfund site, and speficially at Cass Lake, which affects the Leech Lake Band of Objibwe | |
| Gover (2007) | | | Twenty years later- Tribes and the Superfund Program | | | - Describes the inadequate funding allocated to superfund to address tribal needs, and shortcomings in tribal roles in Superfund processes. - Tribal governments should have more opportunities to participate in cleanup and restoration decisions. | |
| Opinion Article | | | | | | | |
| **Author (Year)** | | **Title** | | | **Description** | | |
| Nolan, G (2009) | | Risk Assessment and orphaned/abandoned mines in Canada- What role do aboriginal communities play in risk assessment? | | | - Article written by a chief of the Missanabie Cree First Nation, describing the government plans to address environmental health risks from abandoned mines and highlighting the need to develop policies for working closely with communities, including the integration of communities into initial assessment | | |
| Thesis | | | | | | | |
| **Author (Year)** | | **Title** | | | **Description** | | |
| Clark, Thomas (2020) | | A Comparison of Tribal Sovereignty, Self-Determination, Environmental Justice at the EPA’s Onondaga Lake and Superfund Sites | | | - Explores tribal control over site remediation as an opportunity for alternative and decolonial approaches to remediation and environmental justice within the EPA. | | |

**Table S4:** Studies Measuring Contaminant Concentrations in Environmental Samples

| **Reference** | **Community of Focus, Location** | **Purpose** | **Exposure Studied** | **Analysis Methods** | **Media** | **Results** |
| --- | --- | --- | --- | --- | --- | --- |
| Blake JM, Avasarala S, Artyushkova K, et al. (2015). Elevated Concentrations of U and Co-occurring Metals in Abandoned Mine Wastes in a Northeastern Arizona Native American Community. Environmental Science and Technology. 2015;49(14):8506-8514. | Blue Gap/Tachee Chapter, Navajo Nation, Northeastern Arizona | To assess the presence of metals in the abandoned mine waste site and adjacent springs in the Blue Gap Navajo Chapter | Uranium (U), Arsenic (As), Iron (Fe), Vanadium (V) | Spectroscopy, Microscopy, aqueous Chemistry | Soil (waste solids) | U (6,614 mg kg−1)  V (15,814 mg kg−1)  As (40 mg kg−1)  (all elevated) |
|  |  |  |  |  | Spring Water | U= 67−169 μg L−1  (EPA max 30 μg  L−1) |
| Brown TM, Kuzyk ZZA, Stow JP, et al. (2013). Effects- Based Marine Ecological Risk Assessment at a Polychlorinated Biphenyl-Contaminated Site in Saglek, Labrador, Canada. Environmental Toxicology and Chemistry. 2013;32(2):453-467. | Inukjuak Inuit, Marine Inlets Saglek Bay, Labrador, Canada | Assess Ecological risk of PCB-contaminated marine sediments | Polychlorinated Biphenyls (PCBs) | Sediment toxicity tests (10-d infaunal amphipod survival  [Amphiporeia virginiana] and Microtox [Vibrio fischeri] solidphase)  Biochemistry/physiology measures | Marine Sedmients | Minimum PCB exposure associated with risk to survival or reproduction to sculpin and guillemot is 1000 ng/g wet weight or more, and this occurred within 3 km of the contaminated area for sculpin, and on the site for guillemot |
| Flett L, McLeod CL, McCarty JL, Shaulis BJ, Fain JJ, Krekeler MPS. Monitoring uranium mine pollution on Native American lands: Insights from tree bark particulate matter on the Spokane Reservation, Washington, USA. Environmental Research 2021;194:110619. | Spokane Indian Reservation, Washington | To use Pinus ponderosa as a biomarker to determine the amount of airborne particulate matter associated with the mine | U, Lead (Pb), As, Thorium (Th) | Trace elemental analysis via inductively coupled plasma-mass spectrometry (ICP-MS) | Tree Bark (Ponderosa Pine) | U: 232 ppb  Th: 20 ppb  Pb: 104 ppb  As: 20 ppb  Calculated geoaccumulation index indicate the levels are:  High (U)  Moderate (Th)  Low (Pb, As) |
| Kerfoot WC, Urban N, Jeong J, MacLennan C, Ford S. (2020). Copper-rich “Halo” off Lake Superior's Keweenaw Peninsula and how Mass Mill tailings dispersed onto tribal lands. Journal of Great Lakes Research. 2020;46(5):1423-1443. | L'anse Indian, Lake Superior and Keeweenaw peninsula, Michigan | Examine how mill tailings spread as a dual pulse across southern Keneenaw bay and onto tribal lands | Copper (Cu) | dated sediment cores, beach sand stamps, multi-elemental analyses confirming a tailings origin | Sediment and Mine Tailings | Origin of tailings confirmed to be from the Mill, fluxes in Copper concentrations are elevated |
| Middleton BR, Talaugon S, Young TM, et al. (2019) Bi-directional learning: Identifying contaminants on the Yurok Indian reservation. International Journal of Environmental Research and Public Health. 2019;16(19). | Yurok Tribe,  Lower Klamath, Trinity Rivers, Pacific Coast, California, USA | Identify and address contaminants in the Klamath watershed | Carbamates, dioxins/furans, mercury, microcystins, organochloride pesticides, phenols including PCP and TCP | GC-Q/TOF-MS | Water | Fipronil was the most widely detected target compound  5 target compounds were detected (chlorpyrifos, fipronil, fipronil-sulfide, fipronil-sulfone) |
| Sarkar et. al (2018). Environmental impact assessment of uranium exploration and development on indigenous land in Labrador (Canada): a community-driven initiative | Inuit, Labrador, Canada | To measure the possible  radioactive contamination (total uranium and lead) in  the local ecosystem surrounding an abandoned uranium  development site on indigenous land in Labrador  (Canada). | Uranium and Lead | Inductively coupled mass-spectroscopy | Water, soil, biological samples, tissue samples | Elevated concentrations of U and Pb in stream sediment, Uranium and lead  mobilization in the local environment appears to be  slightly enhanced near the proposed mining site, but  rapidly drops downstream. |

**Table S5:** Studies on contaminants in food

| **Citation** | **Community of Focus, Location** | **Purpose** | **Contaminant Studied** | **Food studied** | **Results** |
| --- | --- | --- | --- | --- | --- |
| Brown TM, Fisk AT, Helbing CC, Reimer KJ. Polychlorinated biphenyl profiles in ringed seals (Pusa Hispida) reveal historical contamination by a military radar station in Labrador, Canada. Environmental Toxicology and Chemistry. 2014;33(3):592-601. | Inukjuak Inuit, Saglek Bay, Labrador, Canada | Assess whether the contaminated site is contributing to the elevated PCB levels in ringed seals from the Northern Labrador coast | PCBs and Organochloride pesticides | Seals | Among 63 ringed  seals sampled along the northern Labrador coast, 5 (8%) had PCB levels that were higher than recorded anywhere else in the Canadian  Arctic. PCB  concentrations in locally contaminated adult males are 2-fold higher than concentrations in those exposed only to long-range PCB  sources and exceed an established threshold of 1.3 mg/kg for adverse health effects in seals. |
| Koch I, Dee J, House K, et al. Bioaccessibility and speciation of arsenic in country foods from contaminated sites in Canada. Science of the Total Environment. 2013;449:1-8. | Indigenous communities in Canada | the objective of the present study is to report the arsenic speciation in bioaccessibility extracts of a number of country foods from contaminated sites in Canada | Arsenic | Hares, edible mushrooms, and wild berries | Berries and plants had lowest bio accessibility, mushrooms and hare meat contained varying amounts of less toxic arsenic |
| Rock T, Camplain R, Teufel-Shone NI, Ingram JC. (2019). Traditional sheep consumption by Navajo people in Cameron, Arizona. *International Journal of Environmental Research and Public Health.* 2019;16(21). | Navajo Nation, Cameron, Arizona | to investigate mutton consumption of the Navajo people living in Cameron | Uranium | Mutton | Mutton is consumed during family events for most chapter members  ceremonies may be altered or threatened due to contamination of mutton |
| Samuel-Nakamura C, Robbins WA, Hodge FS. Uranium and associated heavy metals in Ovis aries in a mining impacted area in northwestern New Mexico. *International Journal of Environmental Research and Public Health.* 2017;14(8). | Navajo Nation Northeastern New Mexico, USA | The objective of this study was to determine heavy metal concentrations in tissue samples collected from sheep (Ovis Aries), the primary meat staple on the Navajo reservation in northwestern New Mexico. | Uranium, As, Cd, Pb, Mo, and Se | Sheep | Reference dietary intake and recommended dietary allowance were exceeded, but tolerable upper limits were not… |
| Schmitt CJ, Brumbaugh WG, Linder GL, Hinck JE. (2006). A screening-level assessment of lead, cadmium, and zinc in fish and crayfish from Northeastern Oklahoma, USA. Environmental Geochemistry and Health. 2006;28(5):445-471. | Quapaw Nation, Northeastern Oklahoma, New Mexico | to evaluate potential human and ecological risks associated with metals in fish and crayfish from mining in the Tri-States Mining District (TSMD) | Lead, Cadmium, Zinc | Fish and Crayfish | concentrations of Pb and Cd in carp, catfish, and crayfish are sufficiently high to represent a potential health risk to human consumers and warrant advisories |
| Garvin EM, Bridge CF, Garvin MS. (2018). Edible wild plants growing in contaminated floodplains: implications for the issuance of tribal consumption advisories within the Grand Lake watershed of northeastern Oklahoma, USA. Environmental Geochemistry and Health. 2018;40(3):999-1025. | 8 tribes and Nations of Northeastern Oklahoma: Eastern Shawnee, Miani, Modoc, Ottawa, Peoria, Quapaw, Seneca-Cayuga, and Wyandotte, Great Lake Watershed | collect and analyze plant species that are commonly consumed by tribes in the floodplain areas previously demonstrated to have elevated soil metal concentrations | Cadmium (Cd), Pb, Zinc (Zn) | Edible Plants (36 species) | Floodplains shown to be a major contamination pathway for metal accumulation in plants, as levels were significantly different than reference samples |

**Table S6:** Studies on Human Health Outcomes

| **Reference** | **Community of Focus, Location** | **Objective** | **Sample size and description** | **Exposure Studied** | **Health**  **Outcome Studied** | **Results** |
| --- | --- | --- | --- | --- | --- | --- |
| Denham M, Schell LM, Deane G, Gallo MV, Ravenscroft J, DeCaprio AP. (2005) Relationship of lead, mercury, mirex, dichlorodiphenyldichloroethylene, hexachlorobenzene, and polychlorinated biphenyls to timing of menarche among Akwesasne Mohawk girls. Pediatrics. 2005;115(2):e127-e134. | Akwesasne Mohawk, St Lawrence River, New York State, USA and Ontario and Quebec, Canada | Examine the relationship between attainment of menarche and levels of 6 environmental pollutants | n= 138 Akwesasne Mohawk girlds | DDE, HCB, PCBs, Mirex, Lead, Mercury | Age at Menarche, Serum Analysis | Mercury- at or below background levels  Lead- Below CDC limit of 10 ug/L  PCB- consistent with cumulative, continuing exposure pattern  Lead associated with a significantly lower probability of having reached menarche, and 4 PCB congeners with a significantly higher probability of having reached menarche. Thus, attainment of menarche may be sensitive to relatively low levels of lead and certain PCBs |
| Fitzgerald, E. F., Brix, K. A., Deres, D. A., Hwang, S. A., Bush, B., Lambert, G., & Tarbell, A. (1996). Polychlorinated biphenyl (PCB) and dichlorodiphenyl dichloroethylene (DDE) exposure among Native American men from contaminated Great Lakes fish and wildlife. Toxicology and industrial health, 12(3-4), 361-368. | Akwesasne Mohawk, St Lawrence River, New York State, USA and Ontario and Quebec, Canada | Investigate the association with consumption of locally caught fish, residential exposure, body burdens of PCBs, and liver enzyme induction | n= 142 Akwesasne men | PCBs, DDE (consumption of fish and residential exposure) | Body burdens of PCBs, liver enzyme induction | No direct relationship between exposure and body burdens, but high potential among mohawk men for occupational and residential exposures based on surveys |
| Goncharov A, Haase RF, Santiago-Rivera A, et al. High serum PCBs are associated with elevation of serum lipids and cardiovascular disease in a Native American Population. Environmental Research. 2008;106(2):226-239. | Akwesasne Mohawk, St Lawrence River, New York State, USA and Ontario and Quebec, Canada | Determine the relationship between contaminant concentrations, serum lipids, and heart disease | n=335 Akwesasne Mohawk Nation members | PCBs | Serum lipids and heart disease | Study supports that there is a relationship between serum PCBs and pesticides and self-reported cardiovascular disease |
| Hund, L., Bedrick, E. J., Miller, C., Huerta, G., Nez, T., Ramone, S., ... & Lewis, J. (2015). A Bayesian framework for estimating disease risk due to exposure to uranium mine and mill waste on the Navajo Nation. Journal of the Royal Statistical Society: Series A (Statistics in Society), 178(4), 1069-1091. | Navajo Nation, Utah, New Mexico, and Arizona, USA | Examine the relationship of Uranium mine waste exposure and kidney disease, diabetes and hypertension | n=1304 Navajo Nation members | Uranium | Kidney disease, diabetes, hypertension | Evidence of associations between chronic diseases and historic mining era and legacy exposures (strongest with acute exposure and kidney disease) |
| Hwang, S. A., Yang, B. Z., Fitzgerald, E. F., Bush, B., & Cook, K. (2001). Fingerprinting PCB patterns among Mohawk women. Journal of Exposure Science & Environmental Epidemiology, 11(3), 184-192. | Akwesasne Mohawk, St Lawrence River, New York State, USA and Ontario and Quebec, Canada | examine the association between contaminated fish consumption and PCB body burden by comparing congener patterns of locally caught fish and breast milk | n=97 Akwesasne Mohawk women | PCBs | Presence of PCBs in breast milk (congener-specific PCB analysis of breast milk) | High PCB burden associated with fish consumption and congener patterns related to yellow perch caught near contaminated site |
| Meltzer G, Avenbuan O, Wu F, et al. (2020). The Ramapough Lunaape Nation: Facing Health Impacts Associated with Proximity to a Superfund Site. Journal of Community Health. 2020;45(6):1196-1204. | Ramapough Lunaape Nation, Ramapo mountains of New Jersey and New York State | Evaluate self-reported exposure to the superfund site in relation to chronic health outcomes | n=151 (97 Ramapough nation members and 54 non-members) | Self-reported exposure measures (i.e.: land use, consumption, proximity) to unspecified toxic dump site wastes including paint sludge | Chronic Diseases (self-reported) | Native americans were 13.84 times more likely than non-natives to face exposure opportunities from the superfund site. Significant association found between bronchitis and exposure. There is a significant association between being a Ringwood Resident of Native American ethnicity, and opportunities for Superfund exposure |
| Fitzgerald, E. F., Hwang, S. A., Langguth, K., Cayo, M., Yang, B. Z., Bush, B., ... & Lauzon, T. (2004). Fish consumption and other environmental exposures and their associations with serum PCB concentrations among Mohawk women at Akwesasne. Environmental research, 94(2), 160-170. | Akwesasne Mohawk, St Lawrence River, New York State, USA and Ontario and Quebec, Canada | To determine whether low rates of local fish consumption from 1990-1992 were sustained among mohawk women who gave birth from 1992-1995. The study also tests the hypothesis that cumulative life exposure to PCBs through fish is associated with serum levels of PCBs in the population, despite current low rates of consumption | N=111 Akwesasne women | PCBs (exposure via soil, air, fish and other foods) | Serum PCBs | Results suggest that although Mohawk women ate relatively large amounts of local fish in the past, they continue to limit their current rates of consumption, and this trend is reflected in their low current body burdens of PCBs. Other exposures are not significantly contributing to serum PCB concentrations |

**Table S7**: Peer-reviewed articles that collected primary qualitative data on the impacts of contaminated sites

| **Reference** | **Community of focus** | **Purpose** | **Methods** | **Results** |
| --- | --- | --- | --- | --- |
| Cassady, J. (2007). A tundra of sickness: the uneasy relationship between toxic waste, TEK, and cultural survival. Arctic Anthropology, 44(1), 87-97. | Inupiak Inuit, Northwest Alaska, USA | To explore environmental, social and moral uncertainties sparked by the discovery of a toxic waste dump, documenting Inupiaq understandings of radiation, transmission, and control of its spread | Ethnographic field work | - The dump impacted all aspects of society - Country foods were still believed to be nutritionally valuable, curative, and preventive - Risk assessments and advisories are not purely scientific and must also be appreciated as cultural, historically situatied, politically charged |
| Hoover E. Cultural and health implications of fish advisories in a Native American community. *Ecological Processes.* 2013;2(4). | Akwesasne Mohawk,  New York State, USA; Ontario and Quebec, Canada | Explore how a Native American community located downstream from a superfund site has been impacted by contamination and the ensuing environmental health studies | Qualitative Interviews with 64 Akwesasne Mohawk members | - Fish advisories have negative physical (diet related) and cultural consequences (loss of language, culture, and social connections attached to fishing) - Economic considerations are given priority over Mohawk health and culture |
| Smith, K., Luginaah, I., & Lockridge, A. (2010). Contaminated’therapeutic landscape: the case of the Aamjiwnaang First Nation in Ontario, Canada. In Geography Research Forum (Vol. 30, pp. 83-102). | Aamjinawing First Nation, Sarnia, Ontario | 1. to explore how the contamination has impacted the residents' perception of the therapeutic nature of the landscape  2. to investigate residents' perceptions of changes in their relationship with mother earth due to contamination | Qualitative Interviews with 18 members of Aamjiwnaang First Nation | - Themes identified from interviews included that contamination creates fear, health impacts are of concern, residents received a lack of clear information, and interviewees felt that their relationship to mother earth was changing |
| Teufel-Shone NI, Chief C, Richards JR, et al. (2021). Development of a Culturally Anchored Qualitative Approach to Conduct and Analyze Focus Group Narratives Collected in Dine (Navajo) Communities to Understand the Impacts of the Gold King Mine Spill of 2015. *International Journal of Environmental Research and Public Health.* 2021;18(17). | Dine (Navajo) Nation, San Juan River, Southwestern USA | To document the socio-cultural impacts of the spill | Focus group qualitative interviews (12 focus groups comprised of Navajo members) | - The spill is not a single incident, but a long history of assaults on Navajo people, including other environmental contaminations, relocation, broken treaties and a lack of agency and voice |

**Table S8: Steps to addressing a contaminated site (Canada and US) ^8-12^**

| **Stage** | **CANADA** | **UNITED STATES** |
| --- | --- | --- |
| Identifying a contaminated site for potential contaminants | Identification | Pre-CERCLA Screening |
| Historical review (and initial classification in the US) | Historical Review   - Literature review - Site visit - Interviews with informed persons | Preliminary Assessment   - Historical Review - HRS Score |
| Initial testing for contaminants | Initial Testing | Site Inspection |
| Initial Classification (Canada) | Classification |  |
| More detailed testing | Detailed testing | Expanded site inspection |
|  | Re-classification | Integrated ESI/Remedial Investigation |
| Development of a remediation strategy | Remediation Strategy/risk management | Remedial Design |
| Remediation | Implementation | Remedial Action |
| Testing again | Confirmatory Sampling | Post Construction and Completion   - Operation and maintenance and long term response actions - Institutional controls - Five year reviews - Site deletion from the national priorities list |
| Ongoing monitoring | Long term Monitoring |  |

**Table S9:** Peer-reviewed literature on Health Risk Assessment

| **Reference** | **Community of Focus, Location** | **Purpose** | **Methods** | **Results** |
| --- | --- | --- | --- | --- |
| Doyle JR, Blais JM, Holmes RD, White PA. (2012). A soil ingestion pilot study of a population following a traditional lifestyle typical of rural or wilderness areas. Science of the Total Environment. 2012;424:110-120. | Xeni Gwet’in First Nation, Chilko Watershed, Cariboo forest region, Nemiah Valley, British Colombia Canada | to determine if soil exposure of rural or wilderness communities via the ingestion pathway is greater than the ingestion values developed for the population at large, which have been used to underpin HHRAs and regulatory decisions pertaining to contaminated sites | Pilot study-mass balance soil ingestion study; to determine mean soil ingestion rates | The resultant soil ingestion rates were higher than estimates currently recommended for HHRAs of adults |
| Harris SG, Harper BL. (1997) A Native American exposure scenario. Risk Analysis. 1997;17(6):789-795. | Umatilla Indian Reservation, Northeastern Oregon, Colombia River Basin | to develop a lifestyle-based subsistence exposure scenario that represents a midrange exposure that a traditional tribal member would receive | Structured Interviews | Community health factors include human health, environmental quality, and socio-cultural quality of life evaluations.  Subsistence scenarios often do not consider less quantifiable risks  Subsistence scenarios risk underestimating actual individual exposures (variable across age, religious practices, non-food exposures) |

**Table S10:** Peer-reviewed literature using secondary data (i.e.: did not collect primary data (i.e.: reviews, discussions, and case studies))

| **Reference** | **Community of Focus, Location** | **Article Type** | **Key points** |
| --- | --- | --- | --- |
| Ballantine, A. (2017). The river mouth speaks: water quality as storyteller in decolonization of the Port of Tacoma. Water History, 9(1), 45-66. | Pullyap Tribe, Washington | Discussion/ Historical Analysis | - The river mouth was shaped/impacted by colonialism and decolonization processes - The tribe contributed positively to the river’s health by fighting back against the port, city, and polluters |
| Lewis J, Hoover J, MacKenzie D. (2017). Mining and Environmental Health Disparities in Native American Communities. *Current Environmental Health Report.* 2017;4(2):130-141. | Indigenous peoples in the Western USA (Arizona, California, Colorado, Idaho, Montanaa, Nevada, New Mexico, Orgon, South Dakota, Utah, Washington, and Wyoming) | Discussion | - Article describes how management (policies, treaties, and infrastructure) and research converge to create chronic exposure and tribal health concerns - There is a need for more tribal studies, and to incorporate traditional practices into toxicity assessment |
| Moore-Nall A. (2015). The legacy of uranium development on or near indian reservations and health implications rekindling public awareness. *Geosciences (Switzerland).* 2015;5(1):15-29. | Navajo, Southern Ute, Ute Mountain, Hopi, Zuni, Laguna, Acoma, and several other Pueblo nations, Eastern Shoshone and Northern Arapho Nations, The Sioux Nations, The Spokane Nation | Review | - Article aims to promote public awareness of the impact of uranium procurement on Native communities, including a legacy of long-term health effects - There is a need for collaborative funding for projects |
| Sandlos, J., & Keeling, A. (2016). Toxic legacies, slow violence, and environmental injustice at Giant Mine, Northwest Territories. Northern Review, (42), 7-21. | Several First Nations and Metis People in the Northwest Territories | Discussion/Historical Review | - Pollution is examined as more than a technical problem, but as a historical agent of colonial dispossession, alienating Indigenous peoples from their traditional territory - Federal governments attempt to confine contamination to the past, minimizing future responsibilities |
| Sandlos J, Keeling A. Claiming the new north: Development and colonialism at the Pine Point mine, Northwest Territories, Canada. *Environment and History.* 2012;18(1):5-34. | Dene First Nation, Dettah and Ndilo Communities | Discussion/historical Review | - Article discusses the Canada mine site and the changes incurred for Indigenous peoples - Mine closures are a mix of good and bad - Positives to mines include a source of identity and economic opportunity |
| Harper B, Harding A, Harris S, Berger P. (2012). Subsistence Exposure Scenarios for Tribal Applications. Human Ecological Risk Assessment. 2012;18(4):810-831. | Confederated tribes of the umatilla Indian Reservation (CTUIR), Spokane, Washoe, Elem Pomo, Quapaw, Wabanaki | Review | - Develops scenarios intended to capture and describe how the resources are used not only in relation to contaminated sites, but how resources were used prior to contamination and how they will be used again - Scenarios are useful to superfund sites, as they can help to understand lifestyle-related risks |
| Harper BL, Flett B, Harris S, Abeyta C, Kirschner F. (2002)The Spokane Tribe's multipathway subsistence exposure scenario and screening level RME. *Risk Anal.* 2002;22(3):513-526. | Spokane Indian Tribe, Eastern Washington | Case Study | - This article presents portions of a multipathway exposure scenario developed in conjunction with the Spokane Tribal Cultural Resources Program. The scenario serves as the basis for a screening-level reasonable maximum exposure (RME) developed for the Midnite Uranium Mine Superfund site. - The scenario is not generalizable to other tribes - The risk assessment process includes understanding of the mistrust of the federal government and should be considered |
| Arquette M, Cole M, Cook K, et al.(2002) Holistic risk-based environmental decision making: A native perspective.Environmental Health Perspectives. 2002;110(SUPPL. 2):259-264. | Mohawks of Akwesasne, Saint Lawrence River, New York, USA | Risk assessment model | - Aims to create a community-defined model for risk assessment that protect and restore both health and traditional cultural practices - Risk assessments have not generally benefitted Native people, due a lack of indigenous involvement, under-resourced communities, and risk assessors and native people have different definitions of health |
| Holifield R. (2012). Environmental Justice as Recognition and Participation in Risk Assessment: Negotiating and Translating Health Risk at a Superfund Site in Indian Country. *Annals of the Association of American Geographers.* 2012;102(3):591-613. | Leech Lake Band of Ojibwe | Discussion | - Describes the human health risk assessment process at the site - The tribe faced distinct environmental justice issues - EPA policy privileges property owners to assess risk - Cumulative effects of contaminated sites on Indigenous peoples are ignored - Traditional tribal lifeways may be incompatible with traditional risk assessments - Risk assessment is regulatory science, and therefore may lack objectivity |
| Wiseman, C. L., & Gobas, F. A. (2002). Balancing risks in the management of contaminated first nations fisheries. International journal of environmental health research, 12(4), 331-342. | Nuu-shah-nulth people, Ahahminquus community of the Mowachaht tribe.  Powell River area- Silammon people, Northern coast Salish | Retrospective Analysis | - An analysis of canada’s risk management strategies, assessing health risks related to traditional food consumption vs. store bought - Found that the risks are comparable, and suggests that advisories and fisheries closures may be substituting one risk for another |
| Emel J, Krueger R. (2003). Spoken but no heard: The promise of the precautionary principle for natural resource development. *Local Environment.* 2003;8(1):9-25. | Fort Belknap Indian Reservation | Case Study | - The precautionary principle is a conservative risk-management philosophy that encourages action with caution when risk is unclear - This principle changes the weight of traditionally defined “economic development” and “health/environmental protection” in contaminated site management |
| Brugge, D., DeLemos, J. L., & Bui, C. (2007). The Sequoyah Corporation fuels release and the Church Rock spill: unpublicized nuclear releases in American Indian communities. American journal of public health, 97(9), 1595-1600. | Unspecified | Discussion | - Examines 2 case studies of nuclear releases in Native American communities, and critiques the follow-up and attention paid to these cases compared to release events in white areas. - The comparative studies had a similar amount of radiation released, and were reported in peer-reviewed literature (Native American releases primarily in grey literature) |
| Sandlos J, Keeling A. (2016). Aboriginal communities, traditional knowledge, and the environmental legacies of extractive development in Canada. *Extractive Industries and Society.* 2016;3(2):278-287. | Dene First Nation, Dettah and Ndilo Communities | Case Study | - Uses transcripts to show that a remediation project failed to acknowledge that Traditional Knowledge (TK) is not a store house, but is woven with historical memories of development - Dene people advocated for the consideration of legacy impacts of the site, but they were considered “not relevant” - Overall, Traditional Knowledge was not adequately included |
| Sistili, B., Metatawabin, M., Iannucci, G., & Tsuji, L. J. (2006). An Aboriginal perspective on the remediation of mid-Canada radar line sites in the Subarctic: A partnership evaluation. Arctic, 142-154. | Fort Albany First Nation (Cree) | Discussion | - Evaluates a partnership between the First Nation and government to determine the existence of a “true” partnership - Article concludes that the partnership was adequate, using criteria from government documents |

**Table S11:** Peer-reviewed articles focused on risk management strategies

| **Reference** | **Community of Focus, Location** | **Purpose** | **Population** | **Methods** | **Results** |
| --- | --- | --- | --- | --- | --- |
| deLemos, J., Brugge, D., Cajero, M., Downs, M., Durant, J. L., George, C. M., ... & Lewis, J. (2009). Development of risk maps to minimize uranium exposures in the Navajo Churchrock mining district. Environmental Health, 8(1), 1-15. | Churchrock chapter, Navajo Nation, Northwestern New Mexico | Develop GIS-based thematic maps as communication tools to clearly identify high risk exposure areas and offer alternatives to minimize public and ecological health impacts | N=151 Navajo nation members | Production of thematic maps derived from environmental sampling records and public health surveys | Maps showed location and quality of unregulated water resources and identified alternatives  Navajo member working groups predicted that the maps could be useful to community members |
| Bland AD, Kegler MC, Escoffery C, Malcoe LH. Understanding childhood lead poisoning preventive behaviors: the roles of self-efficacy, subjective norms, and perceived benefits. *Preventive Medicine.* 2005;41(1):70-78. | 8 tribes and Nations of Northeastern Oklahoma: Eastern Shawnee, Miani, Modoc, Ottawa, Peoria, Quapaw, Seneca-Cayuga, and Wyandotte | to examine both individual and social influences on four behaviors typically targeted in educational programs to prevent lead poisoning (compared Native American caregivers with white caregivers | n=380 (167 Indigenous participants, 213 white participants) | Cross-sectional caregiver interviews, qualitative and quantitative | No significant differences in preventive behavior found between Native American participants and White participants |
| Kegler, M. C., & Malcoe, L. H. (2004). Results from a lay health advisor intervention to prevent lead poisoning among rural Native American children. American Journal of Public Health, 94(10), 1730-1735. | 8 tribes and nations of northeastern oklahoma: Eastern Shawnee, Miani, Modoc, Ottawa, Peoria, Quapaw, Seneca-Cayuga, and Wyandotte | to assess whether community education provided by lay health advisors through existing Native American social networks was an effective strategy for the primary prevention of lead poisoning behaviors | Participants before intervention (n=331) and after (n=387) | Matched blood sampling for the 9 workers on the site (before and after site exposure) | No significant difference found between the paired samples |
| Kegler MC, Malcoe LH, Fedirko V. Primary prevention of lead poisoning in rural Native American children: behavioral outcomes from a community-based intervention in a former mining region. Fam Community Health. 2010;33(1):32-43. | 8 tribes and nations of northeastern oklahoma: Eastern Shawnee, Miani, Modoc, Ottawa, Peoria, Quapaw, Seneca-Cayuga, and Wyandotte | To test the effectiveness of a community-based lay health advisor intervention for primary prevention of lead poisoning among Native American children who lived in a former mining area | n= 332 (Children-parent pairs) | Blood Lead Levels, Children and Cargiver Interviews | Preventative Behaviors Improved for both Native American and White families |

**Table S12:** Peer-reviewed studies collecting primary data on post-remediation land use

| **Reference** | **Community of Focus, Location** | **Purpose** | **Methods and Study Population** | **Results** |
| --- | --- | --- | --- | --- |
| Burger, J. (2004). Recreational rates and future land-use preferences for four Department of Energy sites: consistency despite demographic and geographical differences. Environmental Research, 95(2), 215-223. | Shoshone-Bannock Tribe, Idaho | determine recreational rates to provide insights into possible exposure; to examine perceptions of general environmental problems; and to examine future land use preferences that could inform and guide remediation and restoration decisions for these lands | Structured Interviews (Semi-quantative data) (n= 324 Shoshone-Bannock Tribal Mambers, 1370 other participants) | found that Shonshone-Bannock Indians have higher fishing, camping, hunting rates, and ranked nuclear material processing very low compared to others |
| Burger J. Study of the future land use of a contaminated site: Preferences versus potential use. *Remediation.* 2004;14(4):97-110. | Aleuts, albequerque, Bernalillo County, New Mexico | Examine future land use preferences for the department of energy's los alamos national laboratory as a function of ethnicity for attendees of the los alamos gun show in new mexico | Structured Interview  (Semi-Quantitative)  n=11 (Native American); n=254 (white); n=63 (hispanic) | Native Americans rated camping and returning the land to Native Americans higher than others (however, they were underrepresented in the sample) |
| LeClerc E, Keeling A. (2015) From cutlines to traplines: Post-industrial land use at the Pine Point mine. *Extractive Industries and Society.* 2015;2(1):7-18. | Dene (Deninu Kue First Nation) and Metis Communities of Fort Resolution, Northwest Territories | Examine post-industrial land use at the abandoned pine point mine in Canada' Northwest Terriotires to broaden understanding of how the environmental and socioeconomic legacies of mining are manifested in nearby Indigenous communities | Semi structured interviews, “map biography”  (Semi-Quantitative) | Major changes in aboriginal land use patterns occurred during the development and operational phases of the pine point mine and were part of a broader transition to a mixed economy.  Contemporary land users create unique combinations of land use and wage employment |

**Table S13**: Peer-reviewed articles that collected primary data Indigenous Inclusion and Collaboration

| **Reference** | **Indigenous community of focus** | **Purpose** | **Methods** | **Results** |
| --- | --- | --- | --- | --- |
| Hoover E. "We're not going to be guinea pigs;" Citizen science and environmental health in a Native American Community. *Journal of Science Communication.* 2016;15(1). | Akwesasne Mohawk,  New York, USA and Ontario and Quebec, Canada | Article examines a collaboration between the community and a research university, including challenges and benefits | Semi-structured Interviews with 64 Akwesasne members | **Community benefits**: information gained, education and job skills, grant money  **Researcher benefits:** access to the community, help of mohawk fieldworkers for recruitment, creation of better results  **Challenges:** time, control over data and mistrust of it’s use, science communication |
| Kegler MC, Rigler J, Ravani MK. (2015). Using network analysis to assess the evolution of organizational collaboration in response to a major environmental health threat. Health Education Research. 2010;25(3):413-424. | 8 Tribes and Nations of Northeastern Oklahoma: Eastern Shawnee, Miani, Modoc, Ottawa, Peoria, Quapaw, Seneca-Cayuga, and Wyandotte | To use network analysis to document the evolution in the Tar creek superfund Site in Northeastern Oklahoma from 1997-2005  21 organizations and 8 tribes were interviewed | Interviews with 21 organizations and 8 tribes | Over time, there was an increase in network density; a reduction in hierarchal structure; an increase in tribal linkages with local, state and federal agencies; and an increase in inter-tribal linkages |

**Table S14:** Number of peer-reviewed articles collecting data per contaminant media

| **Contaminant Media** | **# of Articles** |
| --- | --- |
| Human Biomarkers | 9 |
| Water | 5 |
| Wildlife/Wild Game | 7 |
| Plants | 6 |
| Soil | 4 |
| Sediment | 3 |
| Trees | 1 |
| Mine Tailing | 1 |
| Solid Waste | 1 |
| **TOTAL** | 37 |

**Table S15:** Indigenous Communities of Focus in Peer-reviewed Literature

| **Indigenous community of Focus (Peer-reviewed Literature Sources)** | **Location (in literature)** | **# of articles** |
| --- | --- | --- |
| Navajo Nation | USA | 10 |
| Akwesasne Mohawk | Canada and USA | 8 |
| Quapaw Tribe | USA | 6 |
| Ottawa Tribe (Odawa) | USA | 5 |
| Peoria Tribe | USA | 5 |
| Modoc Nation | USA | 5 |
| Shawnee Tribe | USA | 5 |
| Seneca-Cayuga Nation | USA | 5 |
| Wayandotte Nation | USA | 5 |
| Miami Tribe | USA | 5 |
| Spokane Tribe | USA | 4 |
| Inuit | Canada | 4 |
| Confederated Tribes of the Umatilla Indian Reservation (Cayuse, Umatilla, Walla Walla) | USA | 2 |
| Fort Albany Cree First Nation | Canada | 2 |
| Shoshone-Bannock Indians | USA | 1 |
| Xeni Gwet'in First Nation | Canada | 1 |
| Ute mountain Ute Tribe | USA | 1 |
| Keweenaw Bay (Chippewa) | USA | 1 |
| Ramapough Lunaape Nation | Canada | 1 |
| Leech Lake Ojibwe | USA | 1 |
| Southern Ute Tribe | USA | 1 |
| Métis | Canada | 2 |
| Washoe Tribe | USA | 1 |
| Laguna Pueblo Tribe | USA | 1 |
| Aamjinawing First Nation | Canada | 1 |
| Eastern Shoshone Tribe | USA | 1 |
| Assiniboine (Nakoda) | Canada and USA | 1 |
| Sioux Nations | USA | 1 |
| Cherokee Nation | USA | 1 |
| Gros Ventre Tribe (A'ninin) | USA | 1 |
| Northern Arapho Nation | USA | 1 |
| Wabanaki Confederacy | Canada and USA | 1 |
| Hopi Tribe | USA | 1 |
| Elem Pomo Tribe | USA | 1 |
| Zuni Tribe | USA | 1 |
| Yurok Tribe | USA | 1 |
| Pullyap Tribe | USA | 1 |
| Acoma Pueblo Tribe | USA | 1 |
| Dene (Deninu Kue First Nation) | Canada | 1 |
| Aleuts | USA | 1 |

**Table S16**: Contaminated site processes (Canada and US) ^11,13^

| **Stage** | **CANADA** | **UNITED STATES** |
| --- | --- | --- |
| Identifying a contaminated site for potential contaminants | Identification | Pre-cercle Screening |
| Historical review (and initial classification in the US) | Historical Review   - Literature review - Site visit - Interviews with informed persons | Preliminary Assessment   - Historical Review - HRS Score |
| Initial testing for contaminants | Initial Testing | Site Inspection |
| Initial Classification (Canada) | Classification |  |
| More detailed testing | Detailed testing | Expanded site inspection |
|  | Re-classification | Integrated ESI/Remedial Investigation |
| Development of a remediation strategy | Remediation Strategy/risk management | Remedial Design |
| Remediation | Implementation | Remedial Action |
| Testing again | Confirmatory Sampling | Post Construction and Completion   - Operation and maintenance and long term response actions - Institutional controls - Five year reviews - Site deletion from the national priorities list |
| Ongoing monitoring | Long term Monitoring |  |

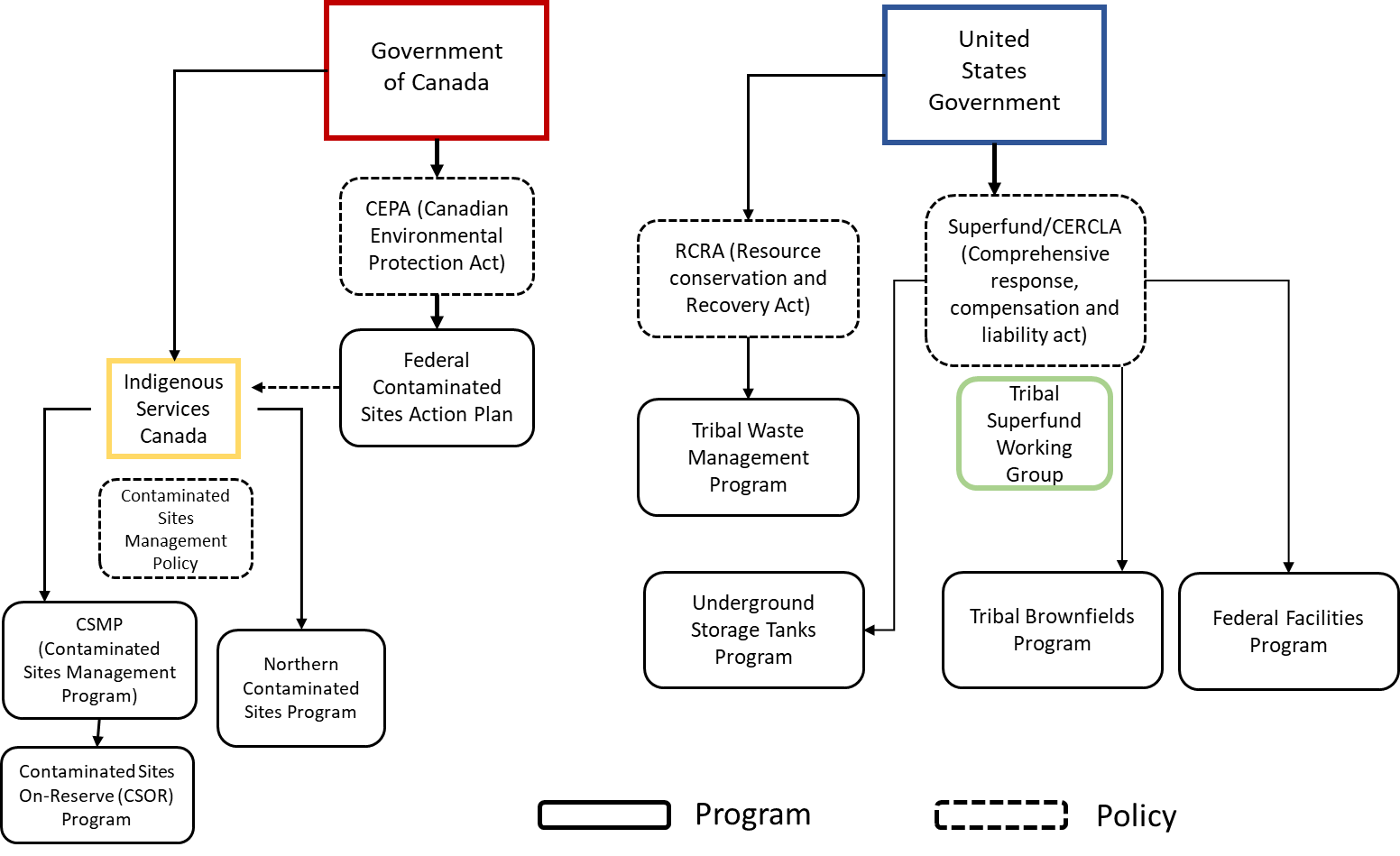

**Figure S1:** Governmental Programming and Legislation Chart ^9-11,13^

**Figure S2: Screenshots of systematic database search**

SCOPUS: Original Search

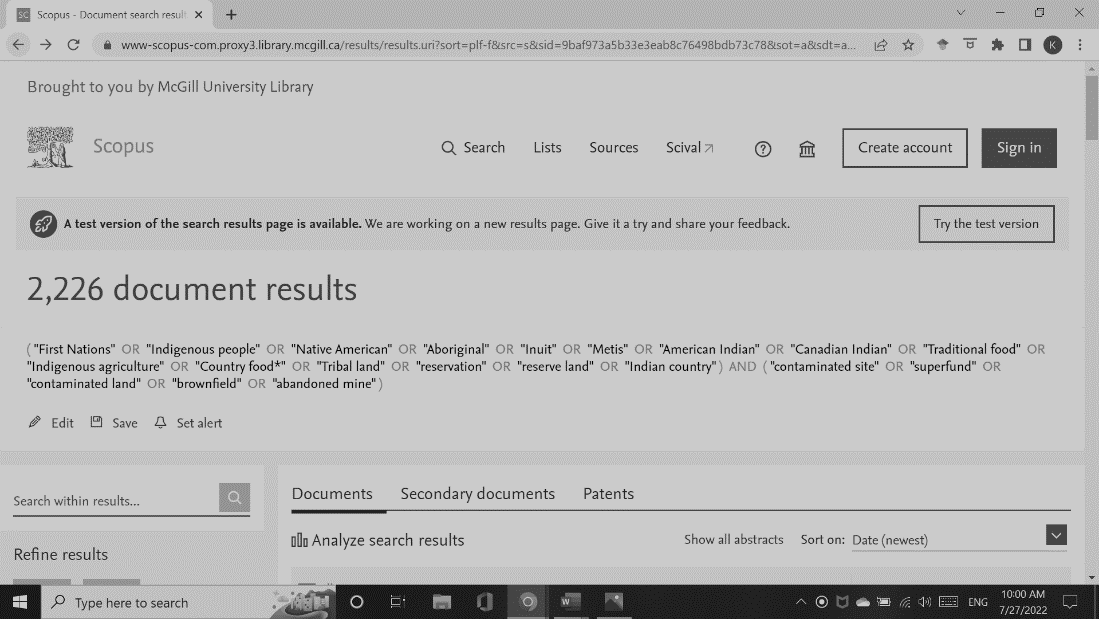

SCOPUS: Search modified to limit to TITLE-ABS-KEY

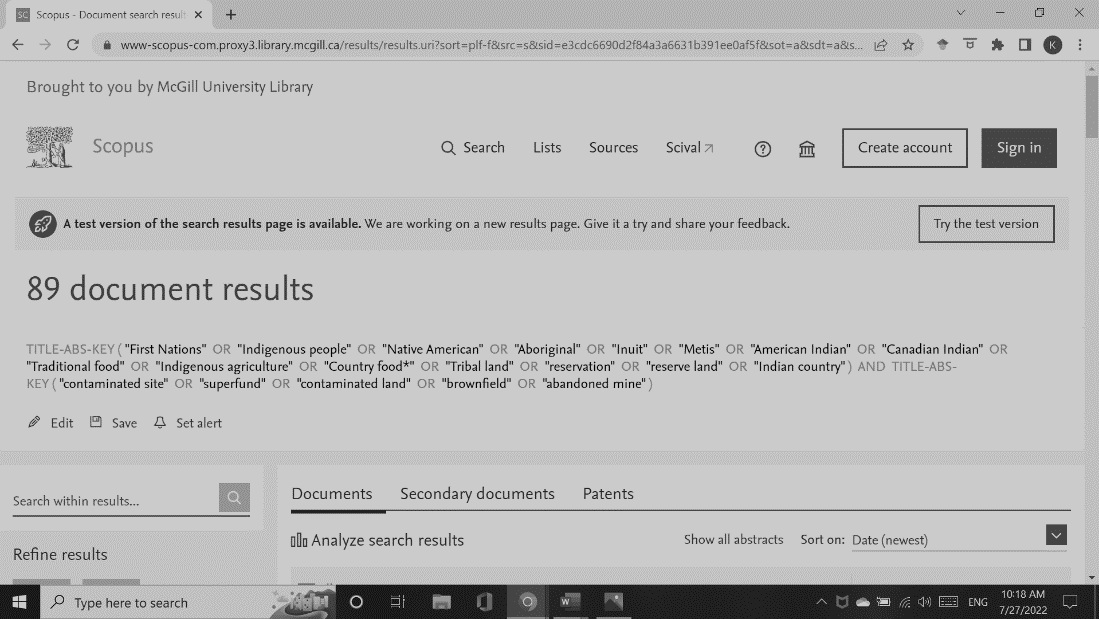

Web of Science Original Search:

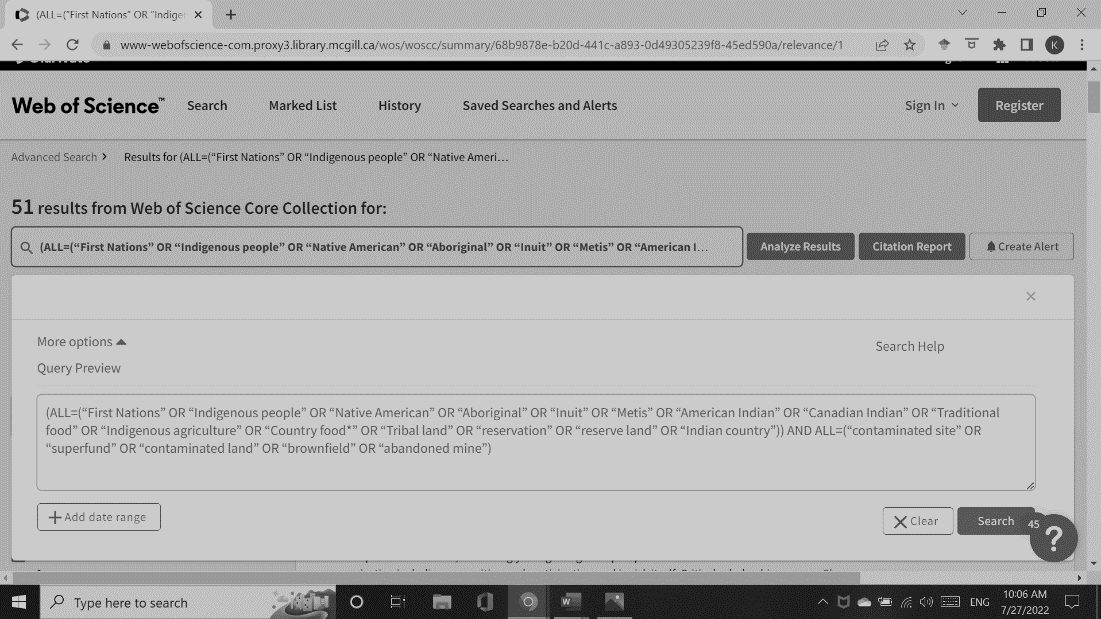

Pubmed Original Search:

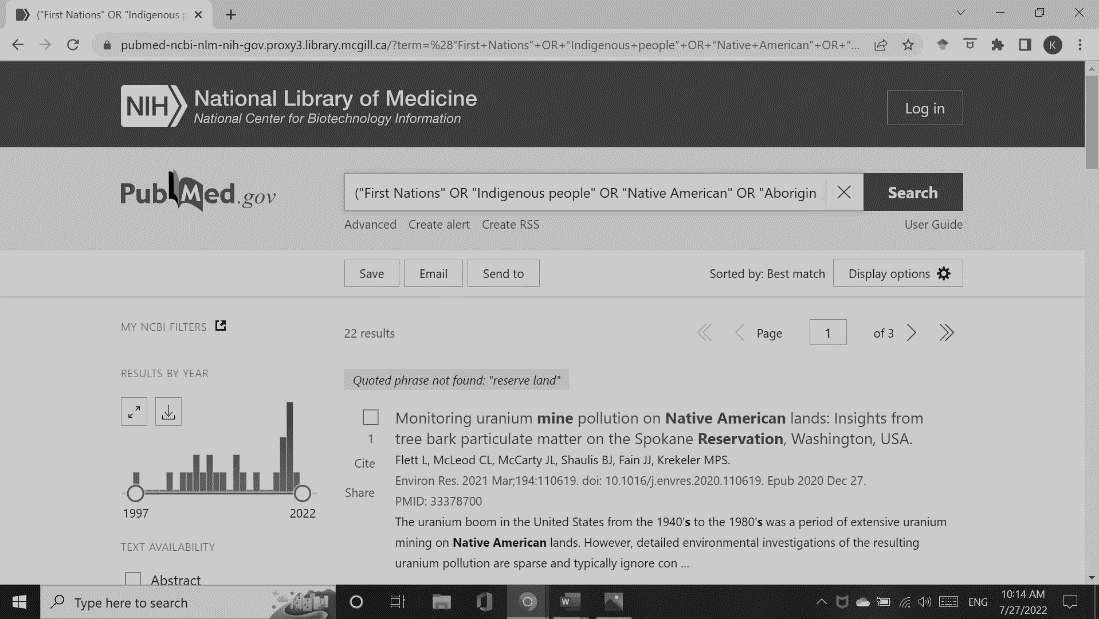

References

1.	Canada Go. Federal Contaminated Sites Inventory. Treasury Board of Canada Secretariat Web site. <https://www.tbs-sct.gc.ca/fcsi-rscf/home-accueil-eng.aspx>. Published 2022. Accessed January 31, 2022.

2.	About Federal Contaminated Sites- Government of Canada. Pollution and waste management Web site. <https://www.canada.ca/en/environment-climate-change/services/federal-contaminated-sites.html>. Published 2022. Accessed2022.

3.	Canada Go. Canadian Environmental Protection Act. In: Canada Go, ed. Ottawa, Ontario 1999.

4.	Action plan for contaminated sites. <https://www.canada.ca/en/environment-climate-change/services/federal-contaminated-sites/action-plan.html>. Published 2021. Updated 2021-04-09. Accessed January 31, 2022, 2022.

5.	EPA. Superfund Glossary. <https://www.epa.gov/superfund/superfund-glossary>. Published 2022. Accessed June 10, 2022.

6.	Bill S-5, Strengthening environmental protection for a healthier Canada act- Summary of Amendments. In: Environment, ed. Ottawa, Ontario 2022.

7.	EPA. Superfund: CERCLA Overview. <https://www.epa.gov/superfund/superfund-cercla-overview>. Published 2022. Accessed January 31, 2022.

8.	Health Canada. *A Guide to Involving Aboriginal Peoples in Contaminated Sites Managament.* Ottawa, Ontario: Health Canada;2010.

9.	Environment Canada. *Federal Contaminated Sites Action Plan (FCSAP) Guidance for Site Closure Tool for Federal Contaminated Sites.* Ottawa, Ontario2012.

10.	Government of Canada. Action plan for contaminated sites. Government of Canada. <https://www.canada.ca/en/environment-climate-change/services/federal-contaminated-sites/action-plan.html>. Published 2019. Accessed May 24, 2022.

11.	EPA. *This is Superfund A community Guide to EPA's Superfund Program.* 2011.

12.	EPA. Contaminated Land. United States Environmental Protection Agency. Report on the Environment Web site. <https://www.epa.gov/report-environment/contaminated-land#roe-indicators>. Published 2021. Updated September 28, 2021. Accessed.

13.	Canada Go. A Federal Approach to Contaminated Sites. In: Group CSMW, ed. Ottawa, Ontario: Dillon Consulting Limited; 1999.
